# Phase Separation Contributes to Pathogenicity for Nonsense Mediated Decay-Escaping Variant Alleles

**DOI:** 10.64898/2025.12.01.25341387

**Authors:** Jiaoyang Xu, Jacob Schmidt, Amy Campbell, Tugce Bozkurt-Yozgatli, Iman Egab, Luisa Mestroni, Matthew Taylor, Jennifer E. Posey, Richard A. Gibbs, Eric Boerwinkle, Paul de Vries, Alanna Morrison, S. Stephen Yi, Chad A. Shaw, Claudia M.B. Carvalho, James R. Lupski, Sujatha Jagannathan, Emel Timucin, Zeynep Coban-Akdemir

**Author notes:** Co-first authors (Jiaoyang Xu, Jacob Schmidt). Correspondence (S.J.), (Z.C.-A.).

## Abstract

Premature termination codons (PTCs) usually trigger nonsense-mediated decay (NMD), but PTCs in the final exon or within 50 base pairs of the last exon–exon junction escape surveillance and can yield stable truncated or altered proteins whose disease mechanisms remain poorly defined. Screening pathogenic and likely pathogenic ClinVar variants, we identified 134 autosomal dominant disease genes enriched for NMD-escape frameshift (N=94) or stopgain (N=64) variants. At the gene level, these genes were indistinguishable from length-matched controls in dosage sensitivity, interaction-network centrality and tissue specificity; only the stopgain class showed greater overlap between its NMD-escape regions and protein–protein interaction interfaces (log-odds ratio +1.22). Comparing pathogenic with benign variants in the same genes, pathogenic alleles truncated further upstream and therefore spanned more post-translational modification sites, nuclear localization signals, protein–protein interaction interfaces and disordered regions; after adjusting for truncation position, pathogenic stopgain variants still overlapped interaction interfaces and Pfam domains more often than benign ones. The two classes remodelled the C-terminus differently: pathogenic frameshift alleles generated novel tails that were less positively charged with a higher phase-separating disordered-region fraction in their divergent segment (q = 0.028), whereas pathogenic stopgain alleles removed longer, more disordered and interaction-prone C-terminal sequence, leaving a truncated protein enriched in arginine and positive charge. NMD-escape disease genes were over-represented among neurodevelopmental-disorder risk genes (35 observed versus 25 expected; odds ratio 1.6, P = 1.3 × 10⁻²), and an NMD-escape truncating variant in *RARB* (microphthalmia, syndromic 12; MIM 615524) formed aberrant condensates in cells. Together, these results define NMD-escape alleles as a mechanistically distinct class of dominant disease variants in which remodelling of interaction-prone, phase-separation-competent C-terminal sequence, rather than transcript loss, drives pathogenicity.

## INTRODUCTION

The classification of variants as pathogenic or benign requires multiple sources of information that vary depending on variant type. Efforts to enhance prediction tools should therefore prioritize efforts focused on the types of variants that are most likely to be pathogenic. In ClinVar, approximately 42% of pathogenic variants are protein truncating variants that introduce premature termination codons (PTCs) into messenger RNAs (mRNAs) (PTC-variants), including stopgain and frameshift alleles. The importance of PTC-variants in human disease is further highlighted by their contribution to a wide variety of disease traits including Mendelian conditions (e.g., paroxysmal kinesigenic dyskinesia[1]), complex diseases (e.g., dilated cardiomyopathy[2]), and quantitative traits (e.g., plasma lipid profile[3]).

A PTC-variant’s impact on disease depends on whether its mRNA is degraded by nonsense-mediated decay (NMD), an evolutionarily conserved cell surveillance mechanism. NMD-triggering variants typically lead to loss-of-function, resulting complete loss in homozygotes; haploinsufficiency in heterozygotes. NMD-escape variants may yield truncated and/or altered proteins with new C-terminal tails that can act via diverse molecular mechanisms: dominant-negative/gain-of-function, loss-of-function, or neutrality, depending on the truncated and/or altered C-terminal context. Hence, predicting NMD status is the first step that shapes downstream variant classification. Current prediction tools about NMD outcomes of PTC-variants are generally based on the exon junction complex-dependent model[4–12], which predicts that mammalian NMD degrades faulty transcripts with PTCs upstream of the last 50-55 base pairs of the penultimate exon - the canonical 50-bp rule[13–15].

The American College of Medical Genetics and Genomics guidelines propose a comprehensive approach for predicting the pathogenicity of PTC-variants with a particular emphasis on the very strong criterion PVS1, indicating a PTC-variant in a gene with a known loss-of-function mechanism[16]. The criterion was further refined, recognizing that not all PTC-variants lead to loss-of-function, some being present with dominant-negative/gain-of-function mechanism. For the NMD-escape variants, the importance of investigating a potential alteration in protein level is highlighted, guided by experimental or clinical evidence. In the absence of such evidence, the rules are arbitrary and need further validation and refinement[17].

A growing number of studies have identified a subset of disease genes that are significantly enriched for NMD-escape pathogenic variants (i.e., NMD-escape disease genes). Examples of these genes include: *DVL1* (Robinow syndrome autosomal dominant 2; MIM:616331)[18], *REST* (Fibromatosis gingival 5; MIM: 617626)[19] and *DVL3* (Robinow syndrome autosomal dominant 3; MIM :616894)[20], *MN1* (CEBALID syndrome; MIM: 618774)[21], *SEMA6B* (Epilepsy, progressive myoclonic, 11; MIM:618876)[22], DAGLA (Neuroocular syndrome 2, paroxysmal type; MIM: 168885)[23] and POGZ (White-Sutton syndrome; MIM: 616364)[24]. In addition, identifying genes enriched with NMD-escape alleles in a broad cohort of patients (∼30K) presented with neurodevelopmental disorders led to several molecular diagnoses through identification of 22 novel disease genes[25]. Despite those recent advances in the field, three key knowledge gaps remain. First, there has been no systematic identification of disease genes that are recurrently affected by NMD-escape disease alleles. Second, the shared molecular and structural properties of these genes have not been delineated. Third, the biophysical mechanisms by which NMD-escape contributes to disease pathogenesis remain poorly understood. Here, we address these gaps by systematically identifying autosomal dominant disease genes enriched for NMD-escape stopgain and frameshift variants, comparing them with length-matched control genes, and comparing pathogenic with benign NMD-escape variants within the same genes. We find that the NMD-escape regions of disease genes are enriched for intrinsically disordered and interaction-prone sequence, and that pathogenic variants affect more functional elements than benign variants of the same gene. The two variant classes alter the C-terminus in different ways: pathogenic frameshift alleles produce novel tails with fewer positively charged residues, more segregated charge and higher predicted phase-separation propensity, whereas pathogenic stopgain alleles remove acidic, disordered sequence. NMD-escape disease genes are enriched for neurodevelopmental-disorder risk genes, and an NMD-escape truncating variant in *RARB* are shown to be associated with aberrant condensate formation in cells. Together, these results define NMD-escape alleles as a mechanistically distinct class of dominant disease variants.

## RESULTS

### GENE-LEVEL DETERMINANTS OF NMD-ESCAPE PATHOGENICITY

#### Enrichment of pathogenic variants within predicted NMD-escape regions of dominant disease genes

To identify disease genes that depend on NMD escape, we built the analysis pipeline shown in **Figure 1B**. Starting from all pathogenic and likely pathogenic protein-truncating variants (PTC-variants) in ClinVar, we kept high-confidence GRCh38 autosomal variants with a valid RefSeq transcript and classified each as NMD-triggering or NMD-escaping under the canonical 50-bp rule, annotating stopgain variants with the aenmd tool and resolving the first downstream premature termination codon of +1 and -1 frameshift variants with a custom caller. For every autosomal-dominant disease gene we then tested whether its NMD-escape PTC-variants were enriched within their NMD-escape region beyond the level expected from rare synonymous variants in the same gene, separately for the stopgain and frameshift classes (Fisher’s exact test with Benjamini–Hochberg correction; the +1 and -1 frameshift sub-classes combined by the ACAT method. This procedure identified 134 autosomal dominant disease genes, each linked in OMIM to at least one autosomal dominant trait, significantly enriched for pathogenic or likely pathogenic PTC-variants within their NMD-escape region (**Figure 1A**; **Table S1**). Of these, 94 genes (70.1%) were enriched for frameshift variants and 64 (47.8%) for stopgain variants, and 24 (17.9%) for both classes.

**Figure 1.**
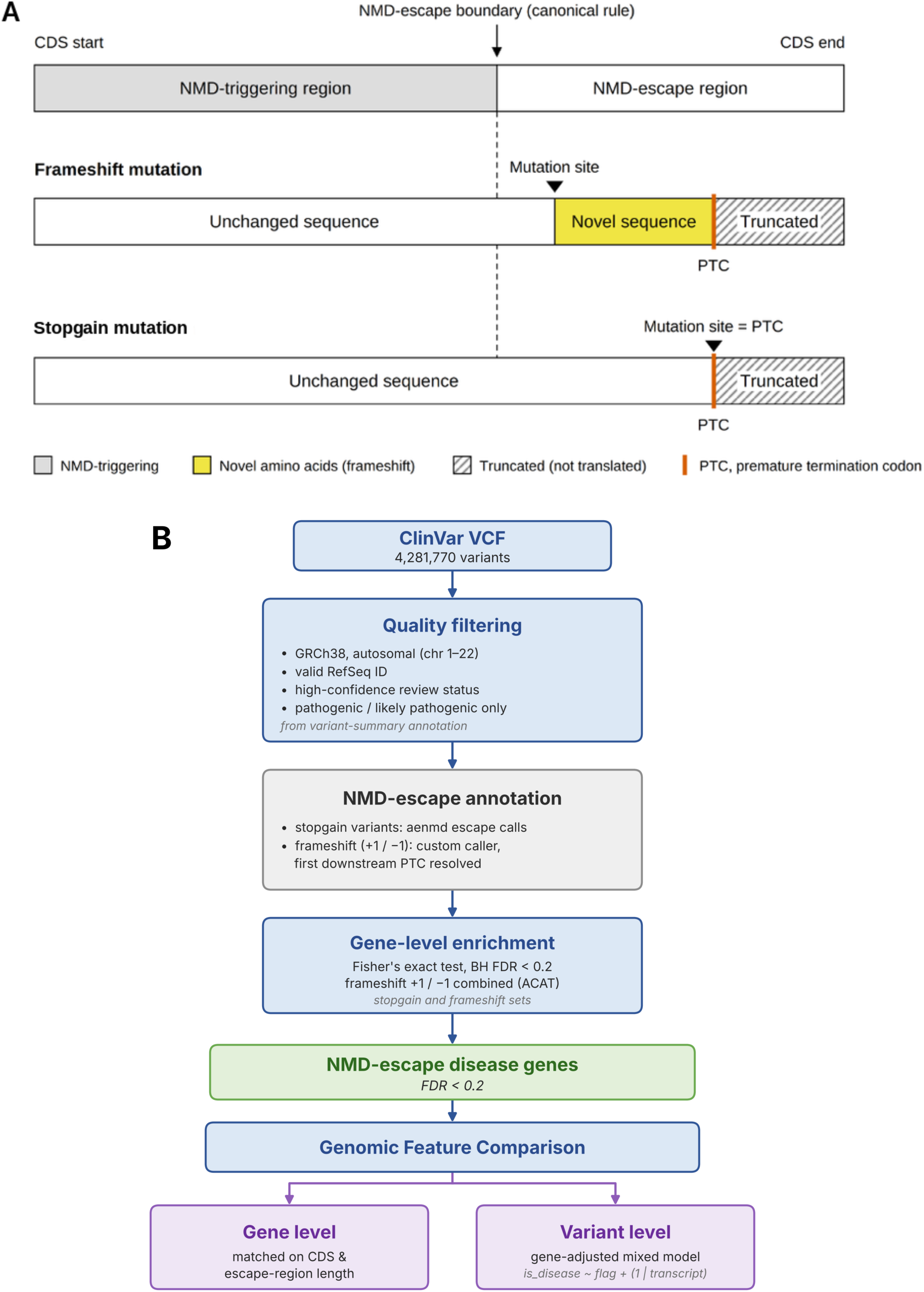
NMD-escape PTC-variant classes and analysis pipeline. **(A)** The canonical 50-bp rule divides the coding sequence into an upstream NMD-triggering region and a downstream NMD-escape region. A frameshift mutation in the escape region shifts the reading frame at the mutation site and adds novel amino-acid sequence up to the first downstream premature termination codon (PTC); a stopgain mutation creates the PTC at the mutation site itself and truncates the protein without adding novel sequence. Hatched segments are not translated. **(B)** Analysis pipeline. Variants from the ClinVar VCF (4,281,770 variants) were quality-filtered (GRCh38; autosomal chromosomes 1–22; valid RefSeq ID; high-confidence review status; pathogenic or likely pathogenic only). NMD escape was annotated with aenmd for stopgain variants and with a custom caller that resolves the first downstream PTC for +1 and -1 frameshift variants, yielding 16,493 NMD-escape pathogenic or likely pathogenic PTC-variants, including 5,266 stopgain variants. Gene-level enrichment was tested separately for stopgain and frameshift variants with Fisher’s exact test and Benjamini–Hochberg correction, combining the +1 and -1 frameshift classes by the ACAT test. The resulting NMD-escape disease genes were compared with length-matched control genes at the gene level (Figures 3 **and 4**) and at the variant level with gene-adjusted mixed models (Figures 5–7). NMD, nonsense-mediated decay; ACAT, aggregated Cauchy association test; VCF, variant call format.

To enable unbiased comparisons throughout the study, we assembled matched control gene sets (**Table S2**). Each control set comprised autosomal dominant disease genes that carry no ClinVar-reported NMD-escape variant but may harbour benign variants available from the control genome aggregation database (gnomAD), genes associated with disease, but not through an NMD-escape–dependent mechanism. This analysis yielded 367 frameshift and 794 stopgain control genes (**Table S2**).

#### Frameshift and stopgain NMD-escape genes are enriched for distinct biological processes

We tested the biological-process enrichment of the NMD-escape disease genes to see which pathways they are enriched for, using the autosomal-dominant disease-gene set as the background (**Figure 2**). The two classes were enriched for distinct biology. Among terms of at least 20 genes ranked by fold enrichment, the frameshift set was dominated by developmental and cell-fate programs: neuron fate commitment (8 of 21 genes, 4.0-fold; *EYA1*, *GATA2*, *NKX2-1*, *NOTCH3*, *SOX9*, *TBR1*, *TGFBR1*, *WNT1*), muscle hypertrophy (7 of 21, 3.5-fold; *CSRP3*, *GATA4*, *MYH7*, *TCAP*), and inner-ear and bone morphogenesis (*EYA1*, *GATA2*, *SOX9*, *TFAP2A*, *WNT1*; *COMP*, *RARB*, *TEK*, *TGFBR2*).

**Figure 2.**
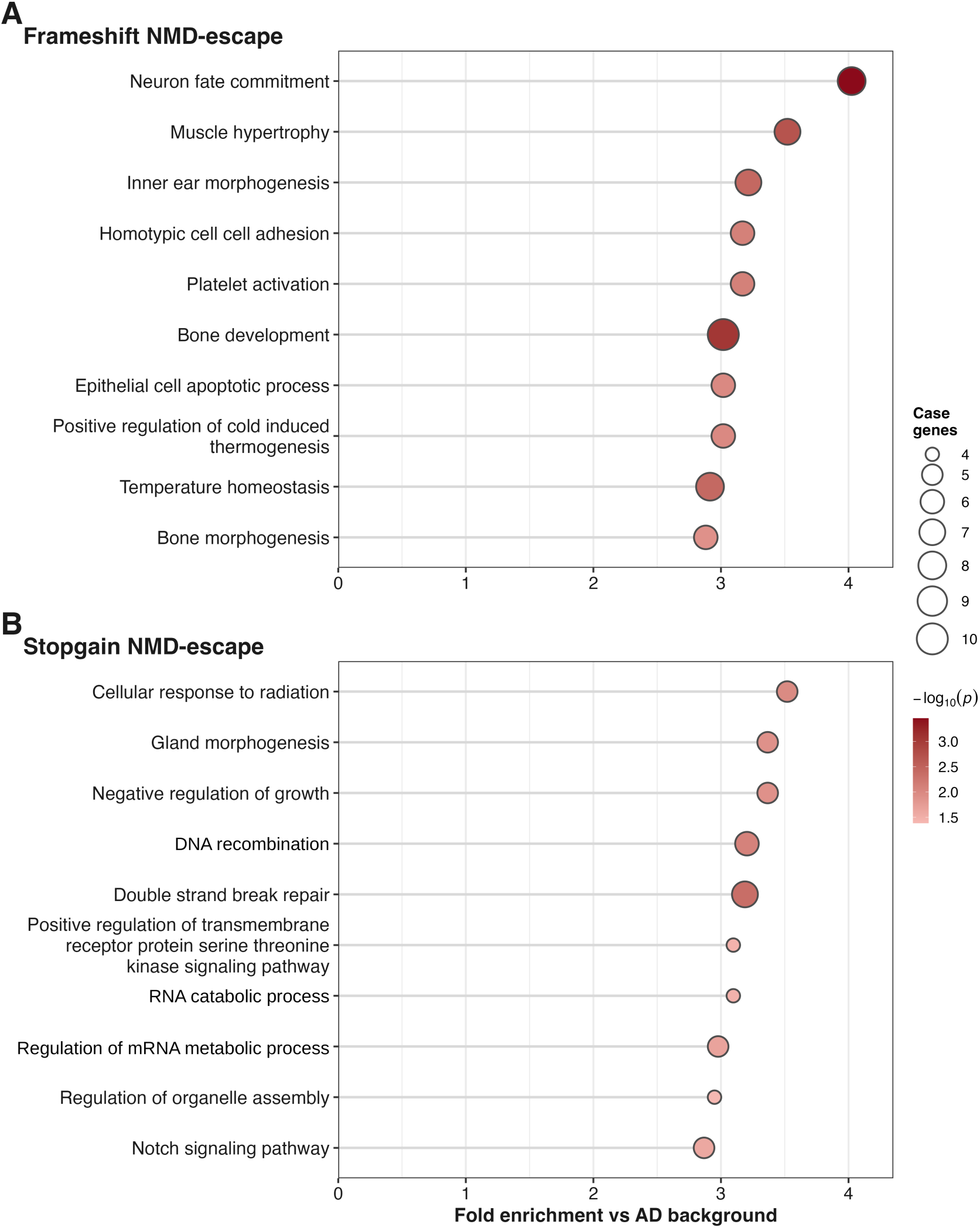
Gene Ontology enrichment of NMD-escape disease genes against an autosomal dominant background. Gene Ontology Biological Process over-representation for each variant class, tested against the full list of autosomal dominant disease genes so that processes common to autosomal dominant genes are not counted as enrichment. Each panel shows the ten terms with the highest fold enrichment among terms of at least 20 genes (a display floor only; all terms of 10–500 genes were tested and enter the Benjamini–Hochberg family). Dot position shows fold enrichment relative to the autosomal dominant background, dot size the number of class genes annotated to the term, and dot colour the significance (-log10 P). **(A)** Frameshift NMD-escape disease genes (94 genes). **(B)** Stopgain NMD-escape disease genes (64 genes). NMD, nonsense-mediated decay.

The stopgain set was instead enriched for genome-maintenance and growth-control processes: double-strand-break repair (7 of 34, 3.2-fold; *BRCA1*, *FUS*, *MLH1*, *PALB2*, *POGZ*, *SMARCD1*, *YY1*), DNA recombination (*BRCA1*, *FUS*, *MLH1*, *PALB2*) and negative regulation of growth (*ACVRL1*, *BRCA1*, *PPP2CA*, *SEMA6B*, *YY1*).

#### NMD-escape disease genes are similar to their length-matched controls in protein–protein interaction degree centrality and tissue specific expression

The case–control design is summarized in **Figure 3A**: within each autosomal-dominant NMD-escape disease gene with pathogenic or likely pathogenic variants extracted from ClinVar as disease variants and with control variants available from the gnomAD database as benign variants. For gene-level comparisons, each disease gene was matched 1:1, without replacement, to a control gene on coding-sequence length and on the ratio of NMD-escape-region length to coding-sequence length. Cases and controls were well matched on these covariates, differing in neither coding-sequence length, NMD-escape-region length nor their ratio (all P > 0.05; **Figure 3B–D**; absolute standardized mean difference < 0.1; **Figure 3E,F**). After adjustment for transcript size, neither protein–protein interaction degree centrality in STRING nor tissue specificity of expression (Tau) differed between disease genes and their controls in either class (paired Wilcoxon signed-rank test; n = 94 frameshift and 64 stopgain pairs; **Figure 3G,H**), indicating that any gene-level pathogenic signal resides instead in the specific protein regions and variants examined below[26].

**Figure 3.**
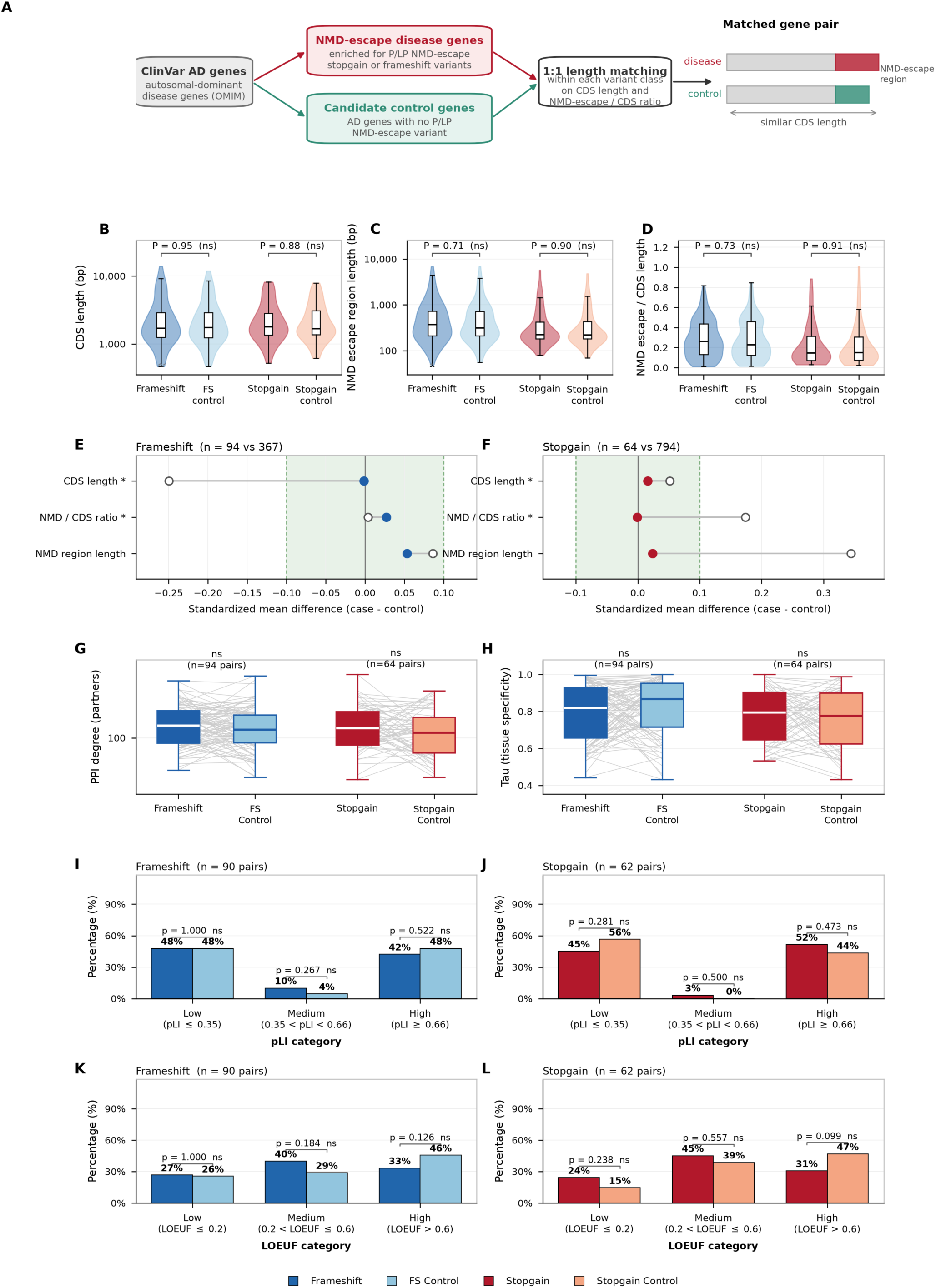
Length-matched control design and gene-level properties of NMD-escape disease genes. **(A)** Selection of control genes. Autosomal dominant (OMIM) disease genes in ClinVar were divided into NMD-escape disease genes, enriched for pathogenic or likely pathogenic NMD-escape stopgain or frameshift variants, and candidate control genes with no pathogenic or likely pathogenic NMD-escape variant. Each disease gene was matched 1:1 to a candidate control gene of the same variant class on coding sequence length and NMD-escape/coding sequence length ratio, and the matched pairs were used for all gene-level comparisons (B–L and Figure 4). **(B–D)** coding sequence length **(B)**, NMD-escape-region length **(C)** and NMD-escape/coding sequence length ratio **(D)** for disease genes and their 1:1 length-matched controls in each class (B and C on a log10 scale). Violins show kernel densities with embedded boxplots (median and IQR); no comparison differs (two-sided Wilcoxon rank-sum test; frameshift/stopgain: coding sequence P = 0.95/0.88, NMD-escape region P = 0.71/0.90, ratio P = 0.73/0.91). **(E, F)** Covariate balance before (open circles) and after (filled circles) matching for frameshift (E; 94 matched pairs from 367 candidate controls) and stopgain (F; 64 pairs from 794 candidates), shown as standardized mean differences (case - control, in case-group standard-deviation units). The shaded band marks absolute standardized mean difference < 0.1, and asterisks mark the matching covariates (coding sequence length and NMD/coding sequence ratio). **(G)** STRING protein–protein interaction degree in matched pairs, which did not differ in either class (paired Wilcoxon signed-rank test; n = 94 frameshift and 64 stopgain pairs). **(H)** Tissue specificity (Tau), not different in either class (n = 94 and 64 pairs). Grey lines connect matched pairs in G and H. **(I–L)** pLI **(I, J)** and LOEUF **(K, L)** categories for disease genes (dark) and matched controls (light) in the frameshift (I, K; n = 94) and stopgain (J, L; n = 64) classes; no category differed between disease genes and controls in either class (all P > 0.05, two-proportion test per category). NMD, nonsense-mediated decay; IQR, interquartile range; pLI, probability of being loss-of-function intolerant; LOEUF, loss-of-function observed/expected upper-bound fraction.

Dosage sensitivity was similarly concordant between groups: the distributions of two gene-level constraint metrics including the probability of loss-of-function intolerance (pLI) and the loss-of-function observed/expected upper-bound fraction (LOEUF) did not differ between NMD-escape disease genes and their matched controls in either class (all P > 0.05; **Figure 3I–L**), indicating that NMD-escape disease genes span a broad range of dosage sensitivity that is well represented among their matched controls.

Nucleotide architecture was also comparable between those genes and their controls. Repeat-element and homopolymer content did not differ between disease genes and controls in either class, whether measured across the full coding sequence, within the NMD-escape region, or as the difference between the two (all P > 0.05; **Figure S1A–F**), and GC content showed no difference across the same three measures (all P > 0.05; **Figure S1G–I**). Nucleotide composition therefore does not distinguish NMD-escape disease genes from their matched controls and is unlikely to account for the protein-level signals described below.

#### Gene redundancy does not distinguish NMD-escape disease genes from their matched controls

NMD-escape variants may yield truncated proteins rather than diminishing their encoded proteins, and are therefore well suited to act through dominant-negative or gain-of-function mechanisms rather than simple haploinsufficiency. A dominant-negative effect generally requires the mutant protein to remain incorporated into multiprotein assemblies, where it can interfere with its wild-type partners, and is less readily tolerated when no redundant gene product can substitute for the affected subunit. We therefore reasoned that, if such mechanisms predominate, NMD-escape disease genes might be preferentially embedded in protein complexes and might carry comparatively little paralogous or isoform redundancy to buffer the perturbation. To test this, we compared gene redundancy indexed by the presence and number of paralogs together with protein-coding-isoform number and membership in curated protein complexes, between the frameshift and stopgain disease gene sets and their length-matched controls.

We observed no such difference. In both classes, 85–90% of disease and control genes had at least one paralog, and the groups differed in the distribution of paralog counts (Mann–Whitney U test, P > 0.05). Within the length-matched framework, paralog number, protein-coding-isoform number and CORUM protein-complex membership were likewise indistinguishable between disease genes and controls in either class (paralogs, frameshift P = 0.1, stopgain P = 0.49; isoforms, frameshift P = 1, stopgain P = 0.93; CORUM complexes, frameshift P = 0.66, stopgain P = 1; **Figure S2A–C**). Gene-family redundancy, isoform number and protein-complex membership therefore do not distinguish NMD-escape disease genes from their matched controls; any dominant-negative contribution is unlikely to be captured by global complex participation and more probably acts through the specific interaction interfaces retained in the truncated protein, which we examine will next.

#### NMD-escape regions of disease genes are enriched for protein-interaction features and intrinsically disordered regions

The NMD-escape regions of disease genes and their matched controls were comparable in length (**Figure 3C**) but differed in feature content. For each matched pair, the escape region of the disease gene and that of its control were scored for the presence of each feature, and the resulting binary flags were compared in a conditional logistic regression stratified by pair (is_disease ∼ flag + strata(pair_id); **Figure 4A,B**). After Benjamini–Hochberg correction, a single feature was enriched in disease-gene escape regions: overlap with protein–protein interaction interface residues, which was elevated in the stopgain class (log-odds ratio +1.22, q = 0.007; 27 discordant pairs in which only the disease gene overlapped an interaction interface versus 8 in which only the control did) but not in the frameshift class. Overlap with a Pfam domain, post-translational modification site, nuclear localization signal, low-complexity sequence, or intrinsically disordered region of at least 20 amino acids did not differ between disease genes and controls in either class (**Figure S3)**. The NMD-escape regions of stopgain disease genes are therefore preferentially enriched for protein–protein interaction interfaces. A truncated protein product encoded by those genes that may still retain the determinants required to assemble into its native complexes, yet lacks an interface encoded downstream of the premature codon. Those truncated products may still be incorporated into those complexes while failing to make the corresponding contact, thereby exerting a potential dominant-negative effect, competing with and sequestering wild-type subunits rather than being eliminated from the cell.

**Figure 4.**
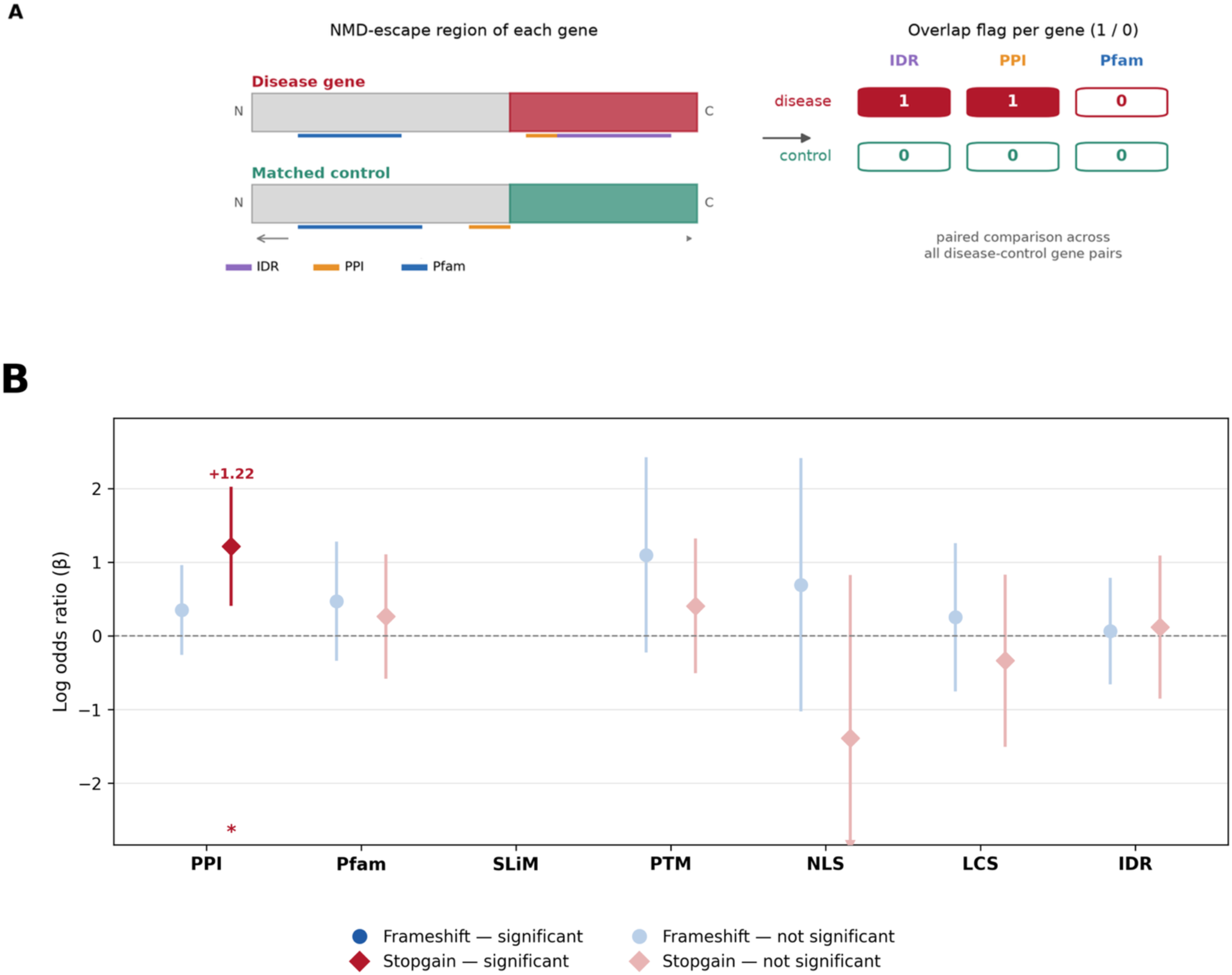
NMD-escape regions of stopgain disease genes are enriched for protein–protein interaction interfaces. **(A)** Design: the NMD-escape region of each disease gene and of its 1:1 length-matched control gene is scored for overlap (1/0) with each feature, and the flags are compared across matched pairs by conditional logistic regression (is_disease ∼ flag + strata(pair_id)), which for a single binary feature is equivalent to McNemar’s test on the discordant pairs. **(B)** Effects were fitted separately for frameshift (circles; up to 94 pairs) and stopgain (diamonds; up to 64 pairs) pairs. Points show the log-odds ratio (β) for carriage of each feature by the disease gene relative to its matched control, and bars the 95% confidence interval. P values are Benjamini– Hochberg-adjusted across the seven features within each class; only the stopgain protein–protein interaction-interface effect is significant (log-odds ratio +1.22, q = 0.007); significant effects (q < 0.05) are shown in dark blue (frameshift) or red (stopgain) and non-significant effects in light shading. Features: protein–protein interaction interface, Pfam domain, short linear motif, post-translational modification site, nuclear localization signal, low-complexity sequence, and intrinsically disordered region (intrinsically disordered region; overlap with a UniProt intrinsically disordered region of at least 20 amino acids). molecular recognition features were excluded because too few genes were informative. short linear motif could not be estimated at the gene level (complete separation in the frameshift class and no discordant pairs in the stopgain class) and is therefore not shown. A downward triangle marks an interval that continues beyond the axis. ***P < 0.001, **P < 0.01, *P < 0.05. NMD, nonsense-mediated decay; β, log-odds ratio.

### VARIANT-LEVEL DETERMINANTS OF NMD-ESCAPE PATHOGENICITY

For variant-level analyses, we compared pathogenic and likely pathogenic NMD-escape variants from ClinVar with population-tolerated (benign) NMD-escape variants from gnomAD in the same disease genes. The dataset comprised 1,318 pathogenic or likely pathogenic and 595 gnomAD frameshift variants, and 413 pathogenic or likely pathogenic and 215 gnomAD stopgain variants (**Table S3**). To account for the uneven distribution of the two variant types across genes, every variant-level comparison was made within genes, using mixed models with gene or transcript as a random effect and fitted separately for the frameshift and stopgain classes.

#### Pathogenic NMD-escape variants truncate and/or alter deeper into functional regions

We next tested whether pathogenic variants affect more functional elements than benign variants of the same gene (**Figure 5A**). For every variant we scored whether the region from the variant to the C-terminus overlaps each annotated feature and compared pathogenic or likely pathogenic with gnomAD variants in a logistic mixed model with a random intercept per gene (is_disease ∼ flag + (1 | gene)), restricted to genes carrying both variant types (**Figure 5B**). In the frameshift class (70 genes; 1,014 pathogenic or likely pathogenic and 590 gnomAD variants), the downstream region of pathogenic variants more often overlapped protein–protein interaction interfaces and nuclear localization signals (log-odds ratio +0.67 and +0.79; Benjamini–Hochberg-adjusted P ≤ 0.021); in the stopgain class (up to 48 genes; 338 pathogenic or likely pathogenic and 215 gnomAD variants) it overlapped protein–protein interaction interfaces, Pfam domains, post-translational modification sites, nuclear localization signals and intrinsically disordered regions (log-odds ratio +0.64 to +1.35; Benjamini–Hochberg-adjusted P ≤ 0.050). Short linear motif and low-complexity-sequence overlap did not differ in either class. These effects carried a confound, however: within genes, pathogenic variants lay further from the C-terminus than gnomAD variants and so left more downstream sequence, consistent with tolerated truncating variants clustering near the 3′ end. A longer downstream region overlaps more features; we therefore refitted each model with log(1 + residues from the variant to the C-terminus end) as a covariate (**Figure 5C**). After this adjustment the frameshift effects fell away, but pathogenic stopgain variants still overlapped protein–protein interaction interfaces (log-odds ratio +1.43; Benjamini–Hochberg-adjusted P = 1.8 × 10⁻⁴) and Pfam domains (+1.07; Benjamini–Hochberg-adjusted P = 6.3 × 10⁻³) more often than benign variants. Pathogenic NMD-escape variants therefore overlap more functional sequence than benign ones partly because they truncate earlier within the escape region, and pathogenic stopgains additionally remove interaction interfaces and Pfam domains beyond what their truncation position alone predicts.

**Figure 5.**
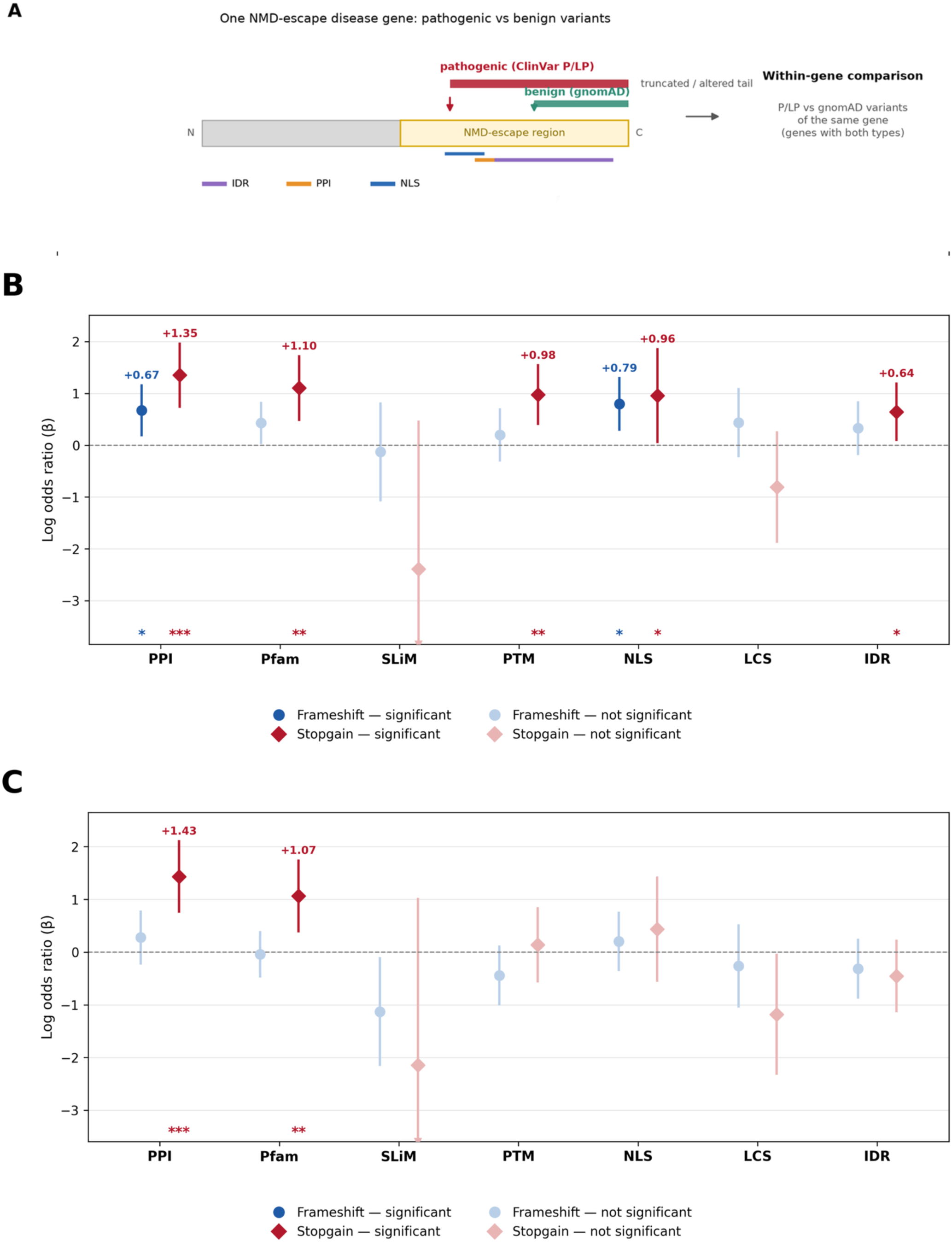
Pathogenic NMD-escape variants truncate further upstream and overlap more functional elements than benign variants of the same gene. **(A)** Design: within each NMD-escape disease gene, the truncated and/or altered tail of each pathogenic or likely pathogenic (ClinVar) and benign (gnomAD) variant, from the variant to the C-terminus, is scored for overlap with each feature, and pathogenic and benign variants of the same gene are compared. **(B, C)** Logistic mixed models with a random intercept per gene, fitted separately for frameshift variants (circles; up to 70 genes, 1,014 pathogenic or likely pathogenic and 590 gnomAD variants) and stopgain variants (diamonds; up to 48 genes, 338 pathogenic or likely pathogenic and 215 gnomAD variants) in genes carrying both pathogenic or likely pathogenic and gnomAD variants of that class. **(B)** Unadjusted model (is_disease ∼ flag + (1 | gene)). **(C)** The same model adjusted for truncation position (is_disease ∼ flag + log(1 + d) + (1 | gene), where d is the number of residues from the variant to the C-terminus); after adjustment, the stopgain protein–protein interaction-interface (log-odds ratio +1.43, q = 1.8 × 10⁻⁴) and Pfam (+1.07, q = 6.3 × 10⁻³) effects remain significant, whereas every other effect, including all frameshift effects, is non-significant. Points show the per-variant log-odds ratio (β) for disease association of each feature, and bars the 95% confidence interval. P values are Benjamini–Hochberg-adjusted across the seven features within each class and panel; significant effects are shown in dark blue (frameshift) or red (stopgain) with β labelled, and non-significant effects in light shading; a downward triangle marks an interval that continues beyond the axis. Features are as in Figure 4, except that Pfam indicates a PTC upstream of the end of the last Pfam domain and intrinsically disordered region indicates at least 20 amino acids of overlap between the region from the variant to the C-terminus end and a UniProt-annotated intrinsically disordered region. ***P < 0.001, **P < 0.01, *P < 0.05. NMD, nonsense-mediated decay; PTC, premature termination codon; β, log-odds ratio.

To make these patterns inspectable at nucleotide and protein resolution, we built a protein-feature viewer that maps nuclear localization signals, Pfam domains, low-complexity sequence and short linear motifs across full-length protein sequences. The viewer lets users compare disease-associated PTC-variants with matched control positions on a per-protein basis, highlighting how interaction and regulatory features cluster in C-terminal regions and how disease variants frequently truncate or alter within or just upstream of these elements (https://github.com/CobanAkdemirlab/NMDescapediseasegene_paper/tree/main/NMDesc-protein-viewer). This resource provides a complementary, protein-centric view that reinforces the aggregate enrichment patterns described above.

#### Pathogenic frameshift tails lose positive charge, whereas pathogenic stopgains remove acidic, disordered sequence

To uncover which features of the altered C-terminus separate pathogenic from benign NMD-escape alleles, we compared pathogenic or likely pathogenic and gnomAD variants of the same transcript with linear mixed models (Δ ∼ cohort + (1 | transcript)), fitted separately for each class. For frameshift variants we modelled the change relative to wild-type in the novel post-frameshift tail (Δ = mutant - wild-type), restricting the analysis to tails of at least 20 amino acids so that composition is estimated reliably (41 shared transcripts; 418 pathogenic or likely pathogenic and 276 gnomAD variants; **Figure 6**). The dominant signal was a loss of positive charge. Among individual amino acids, pathogenic tails gained alanine (q < 0.001) and phenylalanine (q = 0.048) and lost arginine (q = 0.048) relative to benign tails (**Figure 6A**), and the largest class-level shift was gain of nonpolar aliphatic residues (**Figure 6B**). Accordingly, pathogenic tails had lower fractions of basic, positive and R+K residues (all q = 0.003), a lower fraction of charged residues (q = 0.022) and a lower mean net charge (q < 0.001), whereas charge patterning (κ), disorder-promoting fraction, predicted disorder, helix propensity and aromatic content were unchanged, and pLDDT was modestly higher (q = 0.049; **Figure 6C**). Among sequence-composition and charge-patterning proxies of the tail (**Figure 6D**), only sequence charge decoration was lower in pathogenic tails (q < 0.001); the other composition proxies did not differ. Decomposing the charge change localized to the novel out-of-frame tail rather than the wild-type sequence it replaces: although frameshifted tails are intrinsically arginine- and proline-rich, pathogenic variants generated novel tails with significantly less arginine, lower positive charge and more disorder-promoting composition than benign variants of the same transcript, whereas the replaced wild-type C-terminus did not differ (**Figure S4**); gene-level contributions are shown in **Figure S5**. These proxies are composition-based rather than a validated phase-separation predictor; we therefore assessed phase-separation propensity directly with ParSe (**Figure 6E**). Finally, within the divergent segment of the tail, pathogenic tails scored higher on the ParSe phase-separating intrinsically disordered region fraction (q = 0.028) though not on the per-residue classifier score (q = 0.208; **Figure 6E**). Pathogenic frameshift tails are therefore defined by reduced positive charge and, in their divergent segment, a higher predicted phase-separating disordered-region fraction.

**Figure 6.**
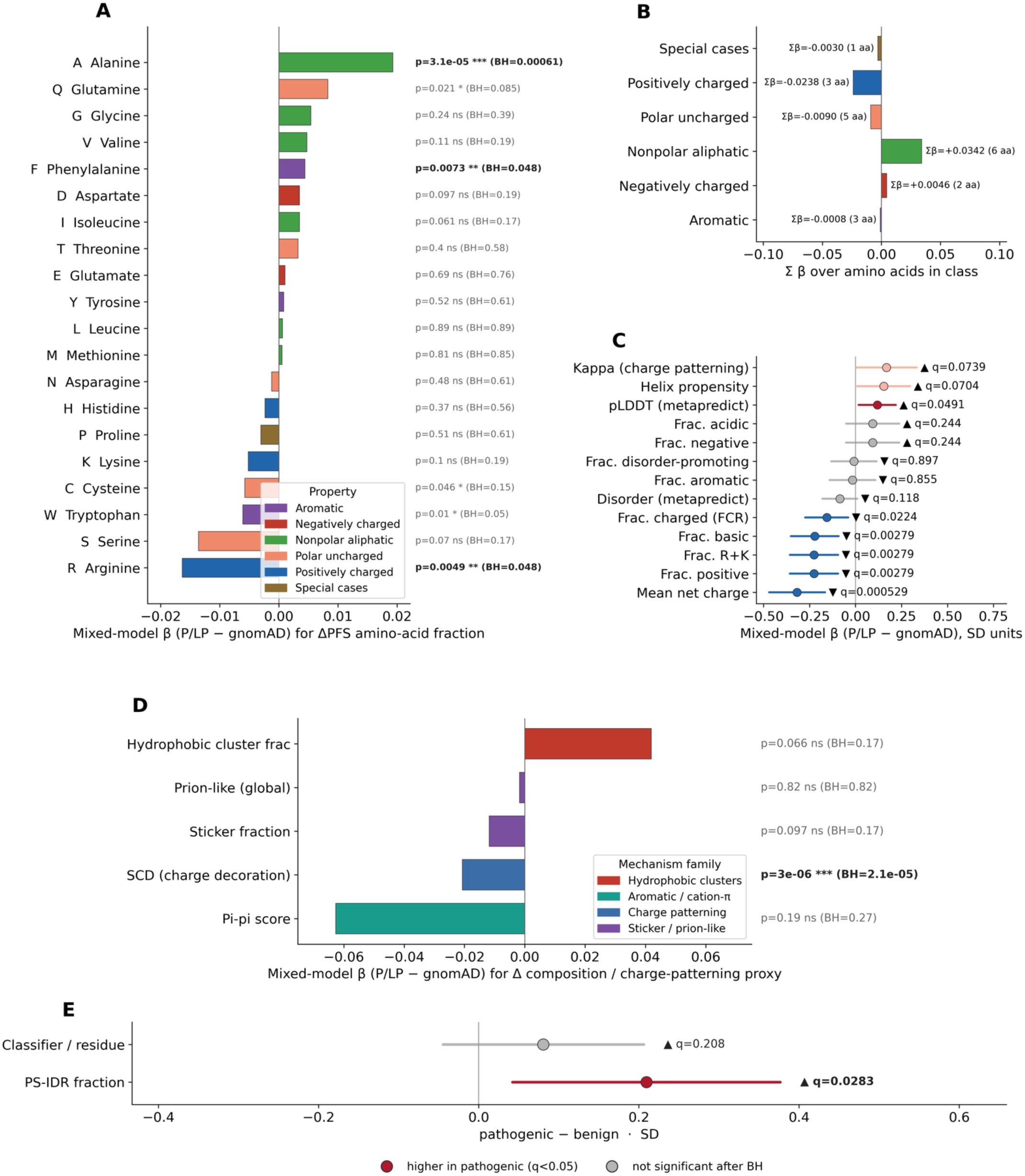
Composition of the post-frameshift tail in pathogenic versus benign NMD-escape frameshift variants. **(A–D)** Pathogenic (pathogenic or likely pathogenic, ClinVar) and benign (gnomAD) frameshift variants of the same transcript with tails of at least 20 amino acids were compared on the change relative to wild-type in the post-frameshift tail (Δ = mutant - wild-type; 41 shared transcripts; 418 pathogenic or likely pathogenic and 276 gnomAD variants) with a linear mixed model per feature (Δ ∼ cohort + (1 | transcript)) and Benjamini– Hochberg correction within each panel. **(A)** Effect (β, pathogenic or likely pathogenic - gnomAD) on the fraction of each amino acid, coloured by physicochemical class; bars to the right of zero mark residues gained more in pathogenic tails. Alanine is higher (q < 0.001) and phenylalanine higher (q = 0.048), while arginine is lower (q = 0.048), in pathogenic tails. **(B)** β summed within each physicochemical class. **(C)** Other tail features (charge metrics, helix propensity, and metapredict disorder and pLDDT) in standard-deviation units: fraction basic, fraction positive and fraction R+K (all q = 0.003), fraction charged (q = 0.022) and mean net charge (q < 0.001) are lower, and pLDDT is modestly higher (q = 0.049), in pathogenic tails; charge patterning (κ), the fraction of disorder-promoting residues, predicted disorder, helix propensity and aromatic content do not differ. **(D)** Effect on sequence-composition and charge-patterning proxies of the tail (prion-like (Q/N/G/S/Y) fraction, Pi-pi score, sticker fraction, sequence charge decoration and hydrophobic-cluster fraction), coloured by mechanism family; sequence charge decoration is lower in pathogenic tails (q < 0.001), while the other proxies do not differ. Charge patterning is captured by two metrics with inverse sign conventions (κ in panel C and sequence charge decoration here), so that a higher κ and a more-negative sequence charge decoration both indicate greater charge segregation. These are composition-based proxies rather than a validated phase-separation predictor; phase-separation propensity itself is shown in panel E. **(E)** The same model applied to the divergent segment of the tail in frameshift variants with both CIDER and ParSe annotations; effects in standard-deviation units (pathogenic - benign) with 95% confidence intervals. The ParSe phase-separating intrinsically disordered region fraction is higher in pathogenic tails (q = 0.028), whereas the per-residue classifier score is not (q = 0.208). In C and E: red, higher in pathogenic; blue, lower; light shading, trend (q < 0.1); grey, not significant. Abbreviations: NMD, nonsense-mediated decay; pLDDT, predicted local distance difference test; κ, the Das–Pappu charge-patterning parameter; R+K, arginine plus lysine; β, mixed-model effect (pathogenic - benign).

Stopgain variants truncate the protein without adding a novel tail, so for this class we compared the truncated mutant proteinwith the full-length wild-type protein (Δ = mutant - wild-type; 47 shared transcripts, 337 pathogenic or likely pathogenic and 210 gnomAD variants; **Figure 7**); the change therefore reflects the composition of the C-terminal segment removed by truncation. Here the dominant signal was the inverse of the frameshift class: a gain of positive charge in the retained protein. The arginine fraction rose (q = 0.003) and the asparagine fraction fell (q = 0.003) more in pathogenic than in benign truncations, with no other amino acid differing after correction (**Figure 7A**); at the class level, the largest shifts were a loss of polar-uncharged residues and a gain of positively charged residues (**Figure 7B**). Consistently, pathogenic stopgains showed higher fractions of basic, positive and R+K residues (q ≤ 0.004), higher charge patterning (κ; q = 0.038) and a higher fraction of charged residues (q = 0.004), together with lower predicted disorder (q < 0.001) and higher pLDDT (q < 0.001), whereas fraction acidic, fraction negative and helix propensity were unchanged (**Figure 7C**). The same composition proxies applied to the whole protein (the entire truncated mutant versus the full-length wild-type; **Figure 7D**) indicated retention of interaction-prone sequence: pathogenic truncations showed a higher Pi-pi score (q = 0.009) and hydrophobic-cluster fraction (q = 0.026), while sticker fraction and sequence charge decoration were unchanged (these remain composition-based proxies, as in the frameshift class). These whole-protein proxies are composition densities: the higher Pi-pi score reflects removal of an arginine-poor, disorder-promoting tail that raises the arginine fraction in the retained protein, rather than a gain of interaction-prone residues within it. Pathogenic stopgains therefore are likelt to remove C-terminal sequence that is more disordered and structurally flexible, leaving a protein enriched in positive charge and in aromatic and hydrophobic, interaction-prone residues. Profiling the removed segments confirmed that the arginine gain reflects loss of long, arginine-poor, polar and disorder-promoting C-terminal sequence rather than removal of arginine-rich regions: pathogenic truncations removed much longer (q < 0.001) and more arginine-depleted (q = 0.002) segments, concentrating arginine in the retained protein (**Figure S6**); gene-level contributions are shown in **Figure S7**.

**Figure 7.**
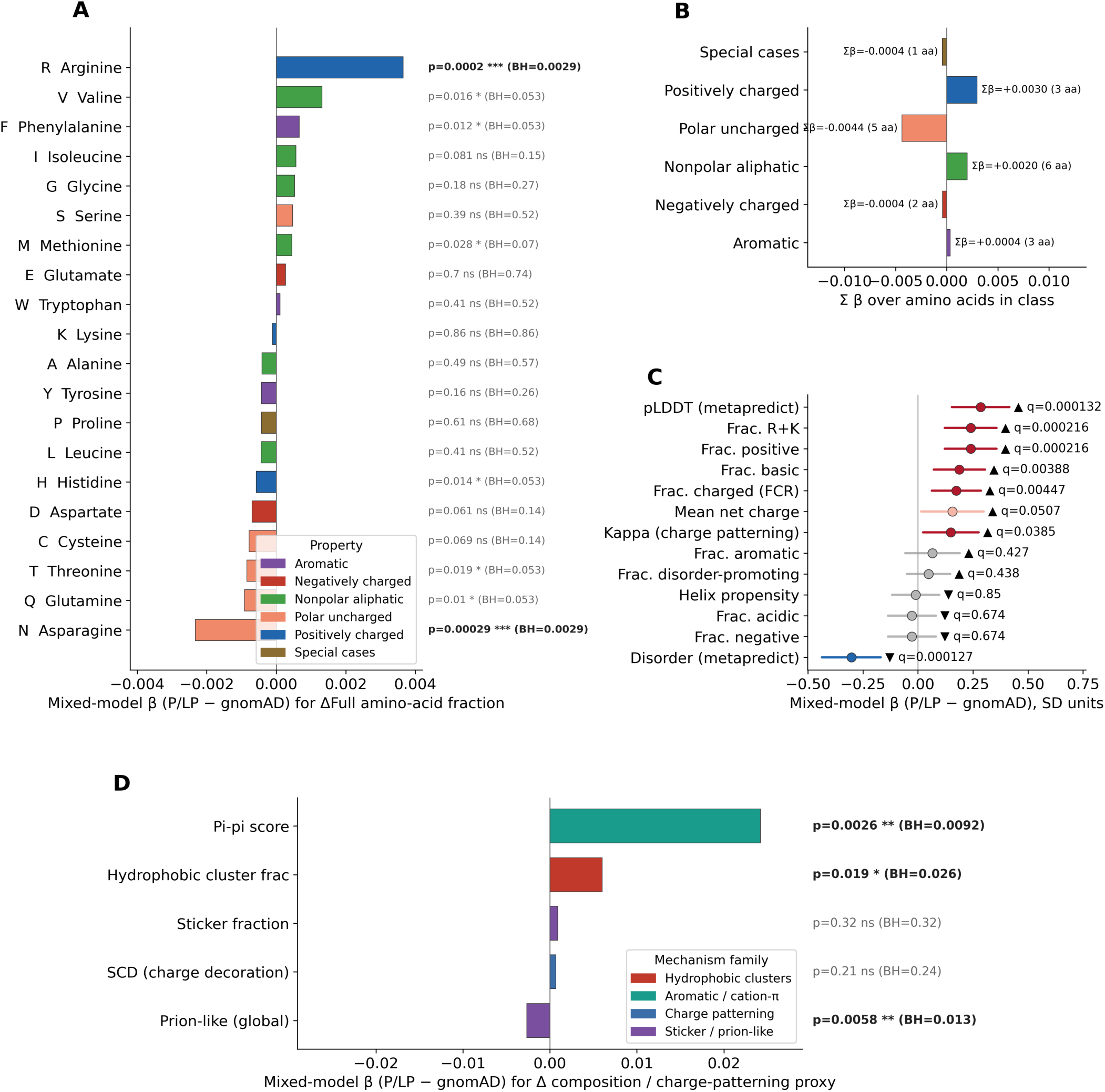
Whole-protein composition change in pathogenic versus benign NMD-escape stopgain variants. Pathogenic (pathogenic or likely pathogenic, ClinVar) and benign (gnomAD) stopgain variants of the same transcript (47 shared transcripts; 337 pathogenic or likely pathogenic and 210 gnomAD variants) were compared on the change between the truncated mutant and the full-length wild-type protein (ΔFull = mutant - wild-type), which reflects the composition of the C-terminal segment removed by the truncation, using a linear mixed model with a random intercept per transcript and Benjamini–Hochberg correction within each panel. **(A)** Effect (β, pathogenic or likely pathogenic - gnomAD) on the fraction of each amino acid, coloured by physicochemical class; bars to the right of zero mark residues whose fraction rises more in pathogenic truncations. Arginine is higher (q = 0.003) and asparagine lower (q = 0.003) after correction. **(B)** β summed within each class; the largest shifts are a loss of polar-uncharged residues and a gain of positively charged residues. **(C)** Other whole-protein features in standard-deviation units: fraction basic, fraction positive and fraction R+K are higher (q ≤ 0.004), fraction charged (q = 0.004) and charge patterning (κ; q = 0.038) are higher, predicted disorder is lower (q < 0.001) and pLDDT higher (q < 0.001) in pathogenic stopgains, whereas fraction negative, fraction acidic and helix propensity do not differ; mean net charge is higher at trend level (q = 0.051). A higher κ indicates more segregated charge patterning. **(D)** Effect on sequence-composition and charge-patterning proxies of the whole protein (prion-like (Q/N/G/S/Y) fraction, Pi-pi score, sticker fraction, sequence charge decoration and hydrophobic-cluster fraction), coloured by mechanism family; the Pi-pi score (q = 0.009) and hydrophobic-cluster fraction (q = 0.026) are higher and the prion-like fraction lower (q = 0.013) in pathogenic truncations, whereas sticker fraction and sequence charge decoration do not differ. These are composition-based proxies rather than a validated phase-separation predictor. Red, higher in pathogenic; blue, lower; light shading, trend; grey, not significant. Abbreviations: NMD, nonsense-mediated decay; pLDDT, predicted local distance difference test; κ, the Das–Pappu charge-patterning parameter; R+K, arginine plus lysine; β, mixed-model effect (pathogenic - benign).

Together, these variant-level analyses show that the two truncating classes are biophysically distinct. Pathogenic frameshift alleles generate novel tails that are depleted of positive charge yet, in their divergent segment, more phase-separation-competent, whereas pathogenic stopgain alleles are more likely to excise disordered, structurally flexible C-terminal sequence, retaining a protein enriched in positive charge and in aromatic, hydrophobic, interaction-prone residues. Despite these opposite directions, pathogenic alleles of both classes showed greater charge patterning than benign alleles: sequence charge decoration was lower in frameshift tails (**Figure 6D**) and κ was higher in stopgains (**Figure 7C**), opposite-signed metrics that both indicate more segregated and blocky charge patterns in mutant proteins.

#### NMD-escape disease genes are enriched for neurodevelopmental disorder risk genes, and a truncating *RARB* variant forms aberrant condensates

We then asked whether NMD-escape disease genes are over-represented among neurodevelopmental-disorder genes. We assembled a curated autosomal dominant risk-gene set from Simons Foundation Autism Research Initiative (SFARI) tier-1 autism genes[27], developmental-disorder genes with definitive evidence in the DECIPHER Developmental Disorder Genotype-to-Phenotype database (DDG2P)[28], developmental and epileptic encephalopathy genes from OMIM[29], and the Epi25 collaborative[30]. Of the 134 NMD-escape disease genes, 35 lay in this combined risk-gene set (**Figure 8A**), significantly more than the 25 expected by chance (odds ratio 1.6, 95% CI 1.1–2.5; P = 1.3 × 10⁻², Fisher’s exact test; **Figure 8B**), linking NMD-escape pathogenic variation specifically to neurodevelopmental disease.

**Figure 8.**
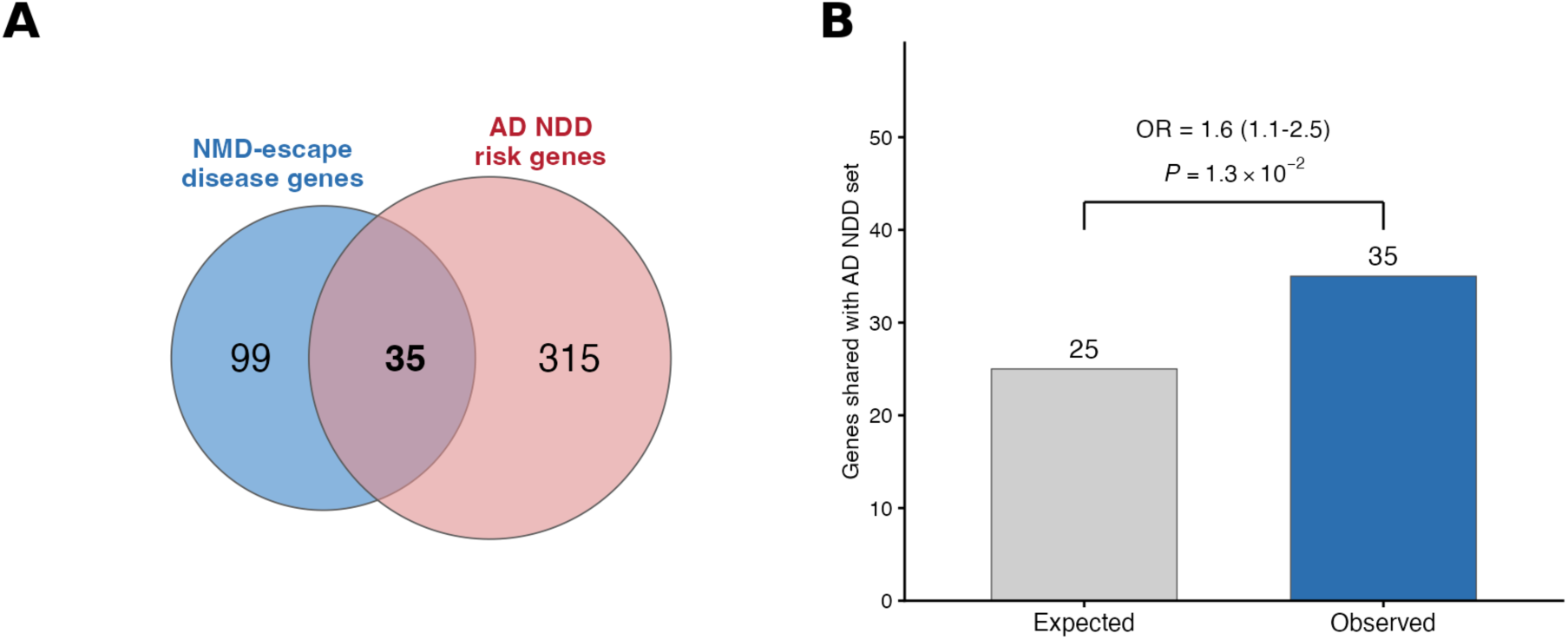
NMD-escape disease genes are enriched for autosomal dominant neurodevelopmental-disorder risk genes. **(A)** Overlap between the 134 NMD-escape disease genes (blue) and 350 autosomal dominant neurodevelopmental-disorder risk genes (pink); 35 genes are shared. **(B)** Number of NMD-escape disease genes shared with the autosomal dominant neurodevelopmental disorder risk-gene set, expected by chance (25) and observed (35); odds ratio 1.6 (95% CI 1.1–2.5), P = 1.3 × 10⁻², Fisher’s exact test. NMD, nonsense-mediated decay; CI, confidence interval.

Many neurodevelopmental-disorder regulators act within biomolecular condensates[31], and the divergent segment of pathogenic frameshift tails showed higher predicted phase-separation propensity than that of benign tails (**Figure 6E**). We therefore examined neurodevelopmental-disorder genes whose proteins carry phase-separation-linked C-terminal intrinsically disordered regions. As a structural illustration, we compared AlphaFold models of wild-type and mutant *RARB* (microphthalmia, syndromic 12; MIM 615524)[32] (**Figure S8**). The variant lies at residue 428, and the two models diverge beyond it: the wild-type C-terminus folds into a short helix, whereas the mutant is truncated and ends in a longer helical segment. APBS electrostatic surfaces (red, negative; blue, positive; white, neutral) show that the mutant introduces lysines at positions that are neutral in the wild-type, creating a prominent positively charged surface patch absent from the wild-type protein, whose corresponding region in wild-type protein is more hydrophobic and neutral (**Figure S8**). These features do not by themselves establish a mechanism, but they are consistent with C-terminal truncation and sequence replacement altering the phase-separation behaviour of *RARB* through its C-terminal intrinsically disordered region (residues 428–454).

To test this prediction experimentally, we expressed RFP-tagged wild-type *RARB* or *RARB* carrying the NMD-escape variant (GRCh38 chr3:25,596,470 C>CCT) in HEK293T cells from a BFP/RFP reporter construct and imaged the protein by fluorescence microscopy (**Figure 9A**). Wild-type *RARB* gave a diffuse RFP signal, whereas the PTC-containing protein formed punctate, non-homogeneous foci (**Figure 9B-C**): fewer than 10% of cells expressing wild-type *RARB* showed a non-homogeneous signal, versus more than 97% of cells expressing the PTC protein (P < 0.0001; **Figure 9D**). Truncating the RARB C-terminus thus markedly changes its distribution in cells, supporting a potetnail pathogenic mechanism based on aberrant condensate formation rather than haploinsufficiency.

**Figure 9.**
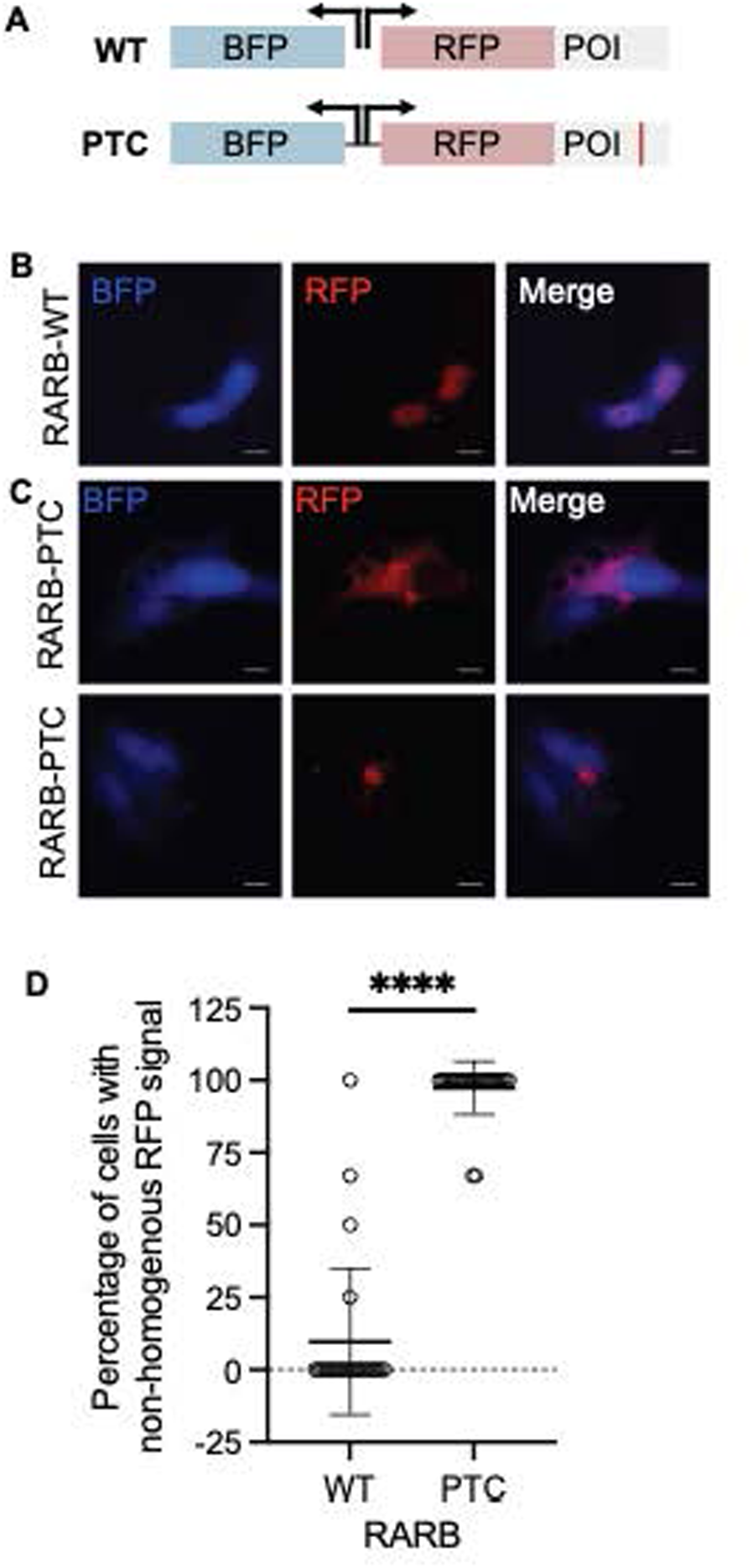
An NMD-escape truncating variant in RARB causes aberrant condensate formation in cells. **(A)** Reporter constructs: BFP and an RFP-tagged protein of interest (POI), either wild-type or carrying the premature termination codon (PTC; red mark), are expressed from the same construct. **(B, C)** Representative fluorescence images of HEK293T cells expressing wild-type **(B)** or PTC-containing (C; two examples) RARB (GRCh38 chr3:25,596,470 C>CCT), showing the BFP, RFP and merged channels. Wild-type RARB gives a diffuse RFP signal, whereas PTC RARB forms punctate, non-homogeneous foci. **(D)** Percentage of BFP⁺/RFP⁺ cells with a non-homogeneous RFP signal for wild-type and PTC RARB (n = 25 images per condition; mean ± s.d.). ****P < 0.0001. NMD, nonsense-mediated decay; s.d., standard deviation.

Together, these results show that NMD-escape disease genes are enriched for neurodevelopmental-disorder risk genes and that an predicted NMD-escape truncation in the *RARB* gene can alter condensate behaviour in cells, pointing to a potential pathogenic mechanism distinct from canonical haploinsufficiency.

## DISCUSSION

Our findings establish NMD-escape PTC-variants as a widespread and mechanistically distinct class of pathogenic alleles in autosomal dominant disease. PTC-variants are traditionally treated as loss-of-function alleles degraded by NMD[33, 34], yet a substantial fraction escape RNA surveillance[35–38]. The resulting stable truncated or altered proteins can perturb interaction networks and condensate organization, mechanisms that fall outside the canonical haploinsufficiency model[39].

By systematically characterizing predicted NMD-escape pathogenic or likely pathogenic variants across 134 autosomal dominant disease genes, we found that at the gene level these genes are indistinguishable from length-matched controls in interaction-network centrality, tissue specific expression and dosage sensitivity, with only the stopgain class showing greater overlap between its NMD-escape regions and protein–protein interaction interface residues. Within the same genes, pathogenic variants truncated further upstream than benign population variants and so spanned more protein–protein interaction interfaces, Pfam domains, post-translational modification sites, nuclear localization signals, and intrinsically disordered regions; after adjusting for truncation position, pathogenic stopgain variants still overlapped interaction interfaces and Pfam domains more often than benign variants, whereas the frameshift signals did not persist. These results support a model in which pathogenic NMD-escape variants remove critical sites in a protein, partly because they lie further from the C-terminus and, for stopgain variants, interaction interfaces and Pfam domains, beyond what truncation position explains, consistent with prior evidence that perturbing highly connected hub proteins disproportionately drives dominant disease[40].

The gene-level comparisons also argue against a simple redundancy explanation. Monogenic disease genes are, genome-wide, enriched for duplicated genes and tend to retain functionally redundant paralogs, a pattern interpreted as redundancy masking deleterious alleles from selection until stochastic differences in paralog expression unmask them [41]. Against this background, NMD-escape disease genes were no more or less paralog-rich than their length-matched autosomal dominant controls and did not differ in protein-coding-isoform number or protein-complex membership. Because our controls are themselves dominant disease-associated genes, this comparison asks the narrower question of whether redundancy further distinguishes NMD-escape disease genes within an already duplication-rich, dominant-gene background, and it does not. The pathogenicity of NMD-escape variants therefore does not appear to be governed by the global redundancy or complex-membership of the affected gene, but by the specific interaction-prone and disordered sequence retained in the truncated protein, consistent with a potential dominant-negative or condensate-related mechanism acting at the level of individual variants rather than whole genes.

Our interface-centred findings also parallel recent analyses of pathogenic missense variation. Dominant-negative missense mutations are strongly enriched at protein–protein interaction interfaces, where a stably folded mutant subunit can poison the assemblies it joins, and gain-of-function and dominant-negative alleles are frequently missed by variant effect predictors trained mainly on loss-of-function signals[42–44]. These studies are based on missense variants, which largely preserve the protein; our results extend the same interface- and mechanism-centred reasoning to NMD-escape truncating variants, which instead remove all sequence downstream of the introduced premature stop codon, underscoring that attention to molecular mechanism can improve the interpretation of protein-truncating as well as missense alleles.

At the sequence level, the two NMD-escape classes behave differently. Pathogenic frameshift tails carried fewer positively charged residues and a lower mean net charge, and, in their divergent segment, were more phase-separation-competent. Both pathogenic and benign frameshift tails were about twice as arginine-rich as the wild-type sequence they replaced (roughly 11–12% arginine versus ∼6.5% in the replaced wild-type), echoing the arginine-rich, basic out-of-frame tails that drive nucleolar mislocalization in the disorders described by Mensah *et al.*[45]; however since benign tails share this feature with pathogenic alleles, arginine-richness alone does not distinguish pathogenic alleles (**Figure S4**). The apparent contrast with that model reflects differences in comparison and gene set: Mensah et al. contrasted each mutant tail with the acidic wild-type tail it replaces (in genes such as *HMGB1*, whose acidic C-terminal disordered regions make the basic-for-acidic swap drive aberrant nucleolar partitioning), whereas we contrast pathogenic with benign frameshift tails of the same genes. The gain of arginine and positive charge relative to wild-type is shared by pathogenic and benign NMD-escape frameshift tails, and pathogenic tails are only marginally less positively charged than benign ones within genes; arginine-richness in our broader set therefore marks frameshift tails in general rather than discriminating pathogenic alleles. We therefore interpret the arginine-rich, nucleolus-partitioning mechanism as operating in a specific subset of genes with acidic disordered C-terminal tails rather than as a universal determinant of pathogenicity. The lower positive charge of pathogenic tails instead arose within the novel out-of-frame sequence itself: within the same transcript, pathogenic frameshifts generated tails with less arginine and a lower fraction of positively charged residues than benign frameshifts (q = 0.003 for both), whereas the replaced wild-type C-terminus did not differ between pathogenic and benign alleles (q = 0.473 and q=0.936) (**Figure S4**). Charge patterning[46] and multivalent interactions among aromatic and other sticker residues[47–50] are key determinants of intrinsically disordered region conformation and condensate formation, so a loss of positive charge in pathogenic tails may alter their interaction and self-association properties rather than simply destabilize the protein. Pathogenic stopgains, by contrast, removed longer and more arginine-depleted C-terminal segments than benign stopgains, leaving a retained protein enriched in arginine and positive charge and more ordered overall (**Figure S6**).

Beyond charge, pathogenic frameshift tails gained alanine (q < 0.001; **Figure 6A**), a residue characteristic of prion-like and alanine-rich low-complexity domains. Alanine-rich sequence is itself a recognized disease mechanism in developmental transcription factors: alanine-repeat expansions in *HOXD13* remodel its transcriptional condensates and cause synpolydactyly[51], and alanine-tract expansions in *ZIC2*, *SOX3*, *ARX* and *PHOX2B* cause neurodevelopmental and craniofacial disorders[52–55]. *ZIC2* and *SOX9*, both key developmental transcription factors whose activity depends on the liquid-liquid phase separation, are among the genes whose pathogenic frameshift tails show the largest gain of Alanine and loss of positive and total charge (**Figure S5**). Alanine is encoded by guanine–cytosine-rich GCN codons, and part of the tail-wide alanine gain may therefore simply reflect the nucleotide composition of the sequence read in the new frame. Together with the lower sequence charge decoration of pathogenic tails representing more blocky and segregated charge distribution and the higher phase-separating disordered-region fraction of their divergent segment (q = 0.028; **Figure 6E**), these compositional changes point to altered condensate behaviour as a plausible consequence of pathogenic frameshift tails.

We also uncovered a link between NMD-escape mechanisms and neurodevelopmental disease: NMD-escape disease genes were enriched for autosomal dominant neurodevelopmental-disorder risk genes (odds ratio 1.6). Dosage-sensitive neurodevelopmental-disorder risk genes frequently encode chromatin regulators and transcription factors that form biomolecular condensates[31, 56, 57], and in HEK293T cells an NMD-escape truncating variant in *RARB* converted the diffuse wild-type protein into punctate foci in nearly all cells. These observations support a model in which some truncated or altered proteins encoded by NMD-escape alleles disrupt the phase-separated compartments that govern transcription, chromatin state or synaptic signalling[45, 58], which may help explain genotype–phenotype relationships at dosage-sensitive neurodevelopmental-disorder loci.

Beyond these mechanistic insights, our findings have direct implications for clinical variant interpretation. Under the American College of Medical Genetics and Genomics framework, NMD outcome is inferred from the canonical 50-bp rule, derived from exon-junction-complex positioning[4–12]: variants with PTCs more than 50– 55 nucleotides upstream of the final exon–exon junction are predicted to trigger NMD, whereas those downstream are predicted to escape. For predicted NMD-escape alleles, the guidelines recommend assessing possible protein-level effects, yet such evidence is rarely available in practice, and many of these variants are therefore retained as variants of uncertain significance[17]. Our results show that the pathogenicity of NMD-escape alleles tracks with quantifiable protein-level features (protein-protein interaction interface, disorder and condensate potential) that could systematically inform reclassification. Incorporating such data-driven features into American College of Medical Genetics and Genomics and ClinGen frameworks could refine PVS1-based rules and reduce ambiguity in PTC-variant interpretation.

In summary, NMD-escape pathogenic and likely pathogenic variants may form a mechanistically distinct and clinically relevant source of pathogenic variation. Their impact may arise not from reduced gene dosage but from structurally and functionally remodelled proteins that may disrupt protein-protein interaction networks and condensate behaviour. Integrating positional NMD rules with protein-feature annotations and phase-separation predictions (and, ultimately, functional validation) will be essential for accurate variant interpretation, particularly in dosage-sensitive and neurodevelopmental-disorder risk genes.

## METHODS

### Study design and overview

We conducted a systematic, gene- and variant-level analysis to identify protein, transcript and biophysical features that distinguish predicted NMD-escape pathogenic or likely pathogenic PTC-variants from benign NMD-escape variants. We analysed two truncating classes, stopgain and frameshift variants (the latter combining -1 and +1 frameshifts), in NMD-escape disease genes associated with autosomal dominant traits. At the gene level, these genes were compared with length-matched control autosomal dominant disease genes that carry no predicted NMD-escape pathogenic or likely pathogenic variants; at the variant level, pathogenic or likely pathogenic variants were compared with benign gnomAD NMD-escape variants in the same genes. Our analyses included (i) NMD outcome prediction, (ii) identification of NMD-escape disease genes enriched with predicted NMD-escape alleles, and iii) characterization of gene-level features of those NMD-escape disease genes relative to matched control disease genes, and iv) characterization of modeling of truncated and/or altered protein sequences and downstream “mutant tails generated by those NMD-escape alleles.

### Variant sources and classification

We identified 171,896 pathogenic or likely pathogenic (pathogenic or likely pathogenic; defined by pathogenic or likely pathogenic in ClinVar clinical significance) PTC-variants available from ClinVar[59] (release 2026-02-01) and 153,668 (96,334 frameshift and 57,334 stopgain) control PTC-variants from gnomAD v4[60]. ClinVar variants were classified as (i) stopgain variants or (ii) frameshift (-1 or+1 frameshift) variants, which are constituted by indels predicted to alter the reading frame and create the first downstream PTC within the shifted frame. Minus1 (-1 frameshift) variants shift the reading frame forward by one nucleotide, resulting from the deletion of one base pair or the insertion of two base pairs. Conversely, plus1 (+1 frameshift) variants shift the reading frame backward by one nucleotide, caused by the insertion of one base pair or the deletion of two base pairs. Variant consequences and transcript mappings were annotated on GRCh38 using Ensembl VEP v106 with dbNSFP[61] and plugin suite. Variants mapping to non-canonical transcripts and variants in single-exon genes were subsequently excluded from downstream analyses.

### NMD outcome prediction

We annotated NMD outcomes of those variants using the canonical 50-bp rule implemented in our annotating escape from nonsense-mediated decay (aenmd) package[62]. Variants were predicted to escape NMD if the variant-induced PTC occurred in the final exon or within 50 base pairs upstream of the final exon–exon junction; all other positions were predicted to trigger NMD. NMD-escape calls for stopgain variants were taken directly from aenmd, while frameshift NMD-escape was annotated with custom code that resolves the first downstream PTC in the shifted reading frame. From the ClinVar variant_summary (downloaded on 02/01/2026) file we retained variants with Assembly = GRCh38, autosomal contigs (chr 1–22), a valid RefSeq ID, a ReviewStatus not containing “no assertion” or “no interpretation,” and a ClinicalSignificance not containing “not provided,” “drug response,” “other,” “risk,” “low penetrance,” “conflicting,” “affects,” “association,” “protective,” or “confers sensitivity”; only variants passing these filters were carried forward. Applying this pipeline to the 4,281,770 ClinVar variants (release 2026-02-01) yielded 235,495 aenmd-annotated variants, of which 171,896 were PTC-variants. Of these, 16,435 were predicted to be NMD-escape based on the canonical 50 bp rule including 5,266 stopgain and 11,169 frameshift variants. After applying the variant_summary filters above, 3,881 stopgain and 8,351 frameshift NMD-escape pathogenic or likely pathogenic variants remained for gene-level analysis.

### OMIM and inheritance pattern

The OMIM genemap2.txt file (downloaded Febrary 01, 2026) was used to identify genes annotated with autosomal dominant inheritance pattern. Disease and control gene sets were filtered to include only genes present in this OMIM autosomal dominant gene list (4,949 genes).

### Disease and control gene sets

For each gene, enrichment of pathogenic or likely pathogenic NMD-escape PTC-variants was assessed with a one-sided Fisher’s exact test (alternative = “greater”), comparing how pathogenic or likely pathogenic PTC-variants partition between the predicted NMD-escape and non-escape regions against the corresponding partition of rare synonymous variants in the same gene, so that each gene’s enrichment is normalized to its own rare-synonymous background as a proxy for local mutation rate and variant ascertainment. P-values were corrected across genes by the Benjamini–Hochberg procedure and disease genes were defined as top 20% from the disease genes with predicted NMD-escape variants. The first downstream PTC generated by +1 and -1 frameshifting variants of the same transcript occurs at closely similar positions; the +1 and -1 frameshift classes were therefore combined when selecting frameshift NMD-escape disease genes, pooling their P-values by the ACAT[63] method. Next, we selected genes from this list that are known to be associated with a disease trait that follow an autosomal dominant inheritance pattern available from the online Mendelian Inheritance in Man (OMIM) database. Control genes were selected from the autosomal dominant genes without any ClinVar pathogenic or likely pathogenic NMD-escape variants but may contain benign NMD-escape variants available from the gnomAD; from this list of genes. Where indicated, we matched or adjusted for gene architecture features (e.g., coding length and NMD-escape region length), constraint metrics including pLI[60] and LOEUF[60, 64] to mitigate ascertainment biases.

### Modeling truncated and/or altered protein sequences

For each predicted NMD-escape variant, we modeled the truncated and/or altered protein. The truncated proteins encoded by stopgain variants were modeled to be truncated at the variant-induced PTC. For frameshifts, we translated the altered sequence *in silico* until the first PTC was encountered in the new reading frame. We exported wild-type and variant protein sequences in FASTA format and delineated the downstream mutant region (“tail”) for amino acid composition and biochemical feature analyses.

### Feature annotation of NMD-escape regions and downstream tails

We mapped interaction- and regulation-associated features onto wild-type sequences and assessed their overlap with (i) NMD-escape regions based on the canonical 50-bp rule and (ii) variant-specific downstream tails. Features included protein-protein interaction interface residues. We mapped UniProt-derived interface residues annotations onto the modeled truncated and/or altered proteins and identified whether the retained NMD-escape regions and variant downstream mutant tails contained interface residues. Overlap between predicted truncated and/or altered sequences in those proteins and interface residues were used as a proxy for potential alteration in protein function. We also annotated post-translational modification sites, PFAM domains, nuclear localization signal motifs, short linear motifs and low complexity sequences (LCSs) available from UniProt[65] and other studies[31, 66–68]. For each feature, its annotated residue positions and the boundaries of the NMD-escape region (gene-level analysis, **Figure 4**) or of the region from the variant to the C-terminus end (variant-level analysis, **Figure 5**) were placed in a common amino-acid coordinate frame, with coding-sequence nucleotide n mapped to residue ⌊(n - 1)/3⌋ + 1, and the region was scored as overlapping the feature (1) when its residue interval intersected at least one annotated residue of that feature on the same protein and as non-overlapping (0) otherwise, with proteins lacking the annotation scored 0. Protein–protein interaction-interface overlap was evaluated at nucleotide resolution, and intrinsically disordered regions were required to overlap the region by at least 20 amino acids. For Pfam, the gene-level flag recorded whether the last Pfam domain extended into the NMD-escape region and the variant-level flag whether the premature termination codon fell upstream of that domain’s end.

### Intrinsically disordered region and structural prediction

We assessed intrinsic disorder with Metapredictor[69, 70] on wild-type and variant proteins, quantifying C-terminal intrinsically disordered regions (≥20 amino acids) that overlapped predicted altered regions. For structure-aware features, we used AlphaFold2 v2.3.2 via ColabFold[71, 72] to generate models for wild-type and variant sequences. Multiple sequence alignments (MSAs) were generated using MMseqs2[73] against the UniRef+ Environmental database[74]. For each variant sequence, five structural models were generated using a single random seed per run. Resulting models were ranked by predicted template modeling-score (pTM-score), and the top-ranked model was further refined via relaxation[75]. Each variant was associated with a corresponding UniProtKB identifier, which was used to retrieve the wild-type structure as reference from the AF2-EBI protein structure database[76]. Variant and wild-type structures were then analyzed using a custom Python script. Structural properties were computed using define secondary structure of proteins (DSSP)[77, 78] to extract per-residue plDDT (predicted local distance difference test) scores and assign secondary structures. In addition to these structural annotations, physicochemical properties including amino acid composition and category-level summaries (aromatics; non-polar aliphatic; polar uncharged; positive/negative; glycine; proline), net charge, aromaticity, and isoelectric point were calculated for each sequence as well. All features were computed both globally and within the NMD-escape region. Summary statistics such as mean, standard deviation and range were calculated for each feature. The resulting dataset allows us to systematically evaluate both global and local structural perturbations associated with predicted NMD-escape pathogenic or likely pathogenic variants and control variants and to unravel shared or context-specific structural changes of NMD-escape variants.

### Network centrality and expression specificity

We computed degree centrality in the STRING protein–protein interaction network and compared disease vs. control genes. Tissue specificity (Tau) was derived from GTEx[79] expression profiles to compare distributional differences between groups. Disease and control gene sets differed in coding sequence length and NMD-escape-region length; we therefore controlled for these covariates through 1:1 matching of each disease gene to a control gene of the same variant class on coding sequence length and NMD-escape-region length, followed by paired testing.

### Constraint, dosage, and paralog content

We annotated pLI[60], LOEUF[64], and dosage sensitivity probabilities (resources and versions in code). Paralog presence and counts were obtained from Ensembl BioMart[80]. We compared the proportion of genes with ≥1 paralog and paralog-count distributions between disease and control sets. Transcript-isoform counts (protein-coding) were obtained from Ensembl BioMart, and protein-complex membership was taken from the CORUM core-complex set, restricted to human complexes with at least two distinct subunits, matching each protein to complexes by both gene symbol and UniProt accession and assigning a count of zero to proteins in no annotated complex. Within the length-matched framework, paralog and protein-coding-isoform counts were compared between each disease gene and its matched control by paired Wilcoxon signed-rank tests; for protein complexes, the fraction of genes belonging to at least one CORUM complex (https://mips.helmholtz-muenchen.de/corum/) was compared between disease genes and controls by the Mann–Whitney U (Wilcoxon rank-sum) test; P-values were Benjamini–Hochberg-adjusted.

### Gene Ontology and pathway enrichment

We performed Gene Ontology enrichment analysis with the R package genekitr for the frameshift and stopgain NMD-escape disease-gene sets. Background gene sets and ontology versions are documented in the GitHub repository. Enrichment P values were corrected by false discovery rate within each analysis. A whole-genome background returns significant terms for almost any set of autosomal dominant disease genes; we therefore repeated the Biological Process enrichment with the full autosomal dominant disease-gene list as the background universe (**Figure 2**).

### Phase separation propensity

We evaluated liquid–liquid phase separation potential with ParSe v2[81], scoring the divergent segment of each frameshift escape tail and the corresponding wild-type sequence and analysing the change relative to wild-type (**Figure 6E**). Sequence-based phase-separation metrics were computed as follows: Q/N/G/S/Y fraction (the fraction of glutamine, asparagine, glycine, serine and tyrosine residues, the residue classes that make up prion-like domains), over the whole tail and in the maximum-scoring 100-residue window; sticker fraction (Y, F, W, R, Q and N); π–π score (aromatic fraction × arginine fraction); hydrophobic-cluster fraction; low-complexity fraction; and sequence charge decoration. These composition- and charge-patterning proxies were computed directly from sequence, not with ParSe, on mutant and wild-type post-frameshift tails for frameshift variants (**Figure 6D**) and on the truncated mutant and full-length wild-type protein for stopgain variants (**Figure 7D**). Pathogenic and benign variants were compared with gene-matched linear mixed models as described under Statistics.

For frameshift variants, amino-acid composition and physicochemical features were computed on the mutant post-frameshift tail and on the wild-type sequence from the same position to the C-terminus; for stopgain variants, they were computed on the truncated mutant and the full-length wild-type protein. Both were analysed as the change relative to wild-type (Δ = mutant - wild-type). Frameshift analyses were restricted to tails of at least 20 amino acids, because composition estimated on very short tails is dominated by sampling noise. Charge patterning was quantified as the Das–Pappu κ parameter[82] using localCIDER v0.1.21: κ = δ/δmax, where δ is the average blob-level charge-asymmetry deviation over sliding blobs of 5 and 6 residues and δmax is δ for the maximally charge-segregated permutation of the same amino-acid composition (κ ∈ [0,1]; 0 = well-mixed charges, 1 = block-segregated). Sequences with no charged residues (δmax = 0) return no value and were excluded; no sequence-length cap was applied. The fraction of charged residues (FCR), fraction of positive and negative residues, fraction of disorder-promoting residues, and mean net charge were computed with the same localCIDER version. Amino-acid composition and the grouped composition fractions (acidic [D+E], basic [R+K+H], R+K, and aromatic [F+Y+W]) were computed directly as residue counts divided by region length. Helix propensity was taken as the predicted α-helix fraction from the ProtParam module of Biopython. Predicted intrinsic disorder (pLDDT) were the mean per-residue values from metapredict. Each feature was tested for a pathogenic-versus-benign difference with a linear mixed model with a random intercept per transcript (Δ ∼ cohort + (1 | transcript)), restricted to transcripts carrying both pathogenic or likely pathogenic and gnomAD variants, with Benjamini–Hochberg correction within each panel; physicochemical features were standardised to standard-deviation units. Sequences and intrinsically disordered region annotations were taken from the variant-level master table after exact VCF-anchor placement of each variant on the Ensembl coding sequence; intrinsically disordered regions were UniProt “Disordered” regions of at least 20 amino acids, mapped onto the Ensembl protein by global alignment where the UniProt canonical isoform differed. As a sensitivity analysis, each model was refitted with a random slope for cohort (Δ ∼ cohort + (1 + cohort | transcript)).

Plasmid construction. Wild-type RARB and RARB carrying the NMD-escape variant **(GRCh38 chr3:25,596,470 C>CCT; RFP-tagged PTC construct pRD-RIPE-RFP-RARB_PTC)** were synthesized as gene fragments (Twist Biosciences) and cloned into the ARTX_ctrl vector (Addgene plasmid #214065) to create RFP-tagged versions; the resulting plasmids are available through Addgene.

#### Cell culture

HEK293T cells were maintained in DMEM (Gibco 11995065) supplemented with 10% Fetal Plus III (Atlas Biologicals FP3-0500-A).

#### Transfections

Cells were transfected in 6-well plates at 70% confluency with 5,000 ng of plasmid DNA using Lipofectamine 2000 (Invitrogen 11668027) according to the manufacturer’s instructions. Thirty hours later, cells were dissociated with TrypLE Express (Gibco 12605010) and seeded onto Poly-D-Lysine-coated coverslips (Gibco A3890401) in new 6-well plates; 24 h later, cells were harvested for fluorescence microscopy.

#### Fluorescence microscopy

Cells were fixed in PBS containing 4% formaldehyde (Thermo Scientific 28908) for 15 min at 37 °C and coverslips were mounted with Fluoromount-G (SouthernBiotech 0100-01). Imaging used a Zeiss Axio Observer 7 Advanced Marianas microscope, an ORCA Fusion BT sCMOS camera, and SlideBook 2023; images were processed and analysed in ImageJ. Aggregation was quantified as the percentage of doubly BFP⁺/RFP⁺ cells (n = 31–64 per condition) with non-homogeneous RFP signal in each image (n = 25 images per condition). Data were plotted as mean ± s.d. in GraphPad Prism 11.

### Statistics

Unless otherwise stated, all tests were two-sided and P < 0.05 was considered statistically significant. Gene-level comparisons used 1:1 length-matched disease–control gene pairs: continuous variables were compared with paired Wilcoxon signed-rank tests, constraint categories with two-proportion tests, and feature flags with linear logistic models with a random intercept per matched pair (is_disease ∼ flag + strata(pair_id). Variant-level comparisons were made within genes, because disease and control variants come from the same genes: all variant-level analyses used logistic mixed models with a random intercept per gene or transcript, restricted to genes carrying both pathogenic or likely pathogenic and gnomAD variants: is_case ∼ flag + (1 | gene) for pathogencity, refitted with log(1 + number of residues from the variant to the C-terminus end) as a covariate to account for truncation position, and Δ ∼ flag+ (1 | transcript) for composition and biophysical features. As a sensitivity analysis, every variant-level model was refitted with an additional random slope for the pathogenic– benign difference (+ (1 + is_case | gene)), which allows this difference to vary between genes; results that did not remain significant in this model are stated in the text. For the variant-level intrinsically disordered region flag, intrinsically disordered regions were UniProt “Disordered” regions of at least 20 amino acids (mapped onto the Ensembl protein by global alignment where the UniProt canonical isoform differed; 127 of 166 proteins carry at least one), and a variant was flagged when the region from the variant to the C-terminus end overlapped an intrinsically disordered region by at least 20 amino acids. P values were Benjamini–Hochberg-adjusted across features within each variant class. For Gene Ontology analyses, enrichment P values were false discovery rate-corrected. Summary statistics are reported as medians and interquartile ranges unless noted otherwise, and exact P values and sample sizes are given in the figure legends and supplementary tables.

## Reproducibility and availability

All code, exact resource versions (e.g., ClinVar, gnomAD[60], STRING[83], UniProt[65], AlphaFold[72, 76]), software parameters, and processed inputs/outputs were deposited in a public repository with a time-stamped DOI. Scripts reproducing every figure and table from raw inputs to final statistics have been provided in our site (https://github.com/CobanAkdemirlab/NMDescapediseasegene_paper).

## Declaration of interests

The authors declare no competing interests.

## Acknowledgments

This work was supported in part by the US National Human Genome Research Institute/National Heart Blood Lung Institute jointly funded Baylor-Hopkins Center for Mendelian Genomics (UM1HG006542), by the National Institutes of Health (NIH) (5R01 HD039056, 5R01 HL091771), by the Genomic Research Elucidates the Genetics of Rare Disease (GREGoR) Program (U01 HG011758) to J.E.P., J.R.L., and R.A.G., and by the National Institute of Neurological Disorders and Stroke (NINDS R35 NS105078) to J.R.L. J.X. was supported by Simons Foundation pilot award (AGT011737). J.S. was supported by the Genomic Research Elucidates the Genetics of Rare Disease (GREGoR) Program (U01 HG011758). S.J was supported by the University of Colorado School of Medicine Translational Research Scholars Program, Simons Foundation pilot award (AGT011737), and the National Institutes of Health grant R35GM133433. Z.C.-A. was supported by the TOPMed NHLBI Fellowship, Simons Foundation pilot award (AGT011737) and the Genomic Research Elucidates the Genetics of Rare Disease (GREGoR) Program (U01 HG011758). S.S.Y. is a Partner Faculty Member of the GREGoR Consortium. The content is solely the responsibility of the authors and does not necessarily represent the official views of the NIH.

## Author contributions

Conceptualization, J.X, J.S, L.M., M.T., J.E.P, R.A.G, E.B., P.D.V, A.M., C.M.B, C.S., M.K.E, J.R.L, S.J, E.T and Z.C.-A.; data acquisition, J.X, J.S, S.J, E.T and Z.C.-A; data analysis, J.X, J.S, T.B.-Y., I.E., C.M.B, C.S., M.K.E, J.R.L, S.J, E.T and Z.C.-A; funding acquisition, J.R.L., J.E.P., R.A.G., E.B., S.J., Z.C.-A.; visualization, J.X, J.S, Z.C.-A.; writing – original draft, J.X, J.S., J.R.L., S.J., E.T. and Z.C.-A..; writing – review and editing, all authors. All co-authors read and approved the final manuscript.

## Data availability

All datasets analyzed in this study are publicly available. ClinVar pathogenic or likely pathogenic PTC-variants (downloaded 08/19/2024), gnomAD v4 population PTC-variants, OMIM genemap2 (downloaded 10/25/2024), and supporting annotations/resources (UniProt, STRING, GTEx, AlphaFold, ParSe v2, and Ensembl/VEP reference files) were accessed from their respective repositories. The processed variant and gene lists, feature matrices underlying all figures and tables, and supplementary outputs generated in this work are provided in the Supplementary Information and in the project repository: https://github.com/CobanAkdemirlab/NMDescapediseasegene_paper.

## Code availability

The code used in this study is available in (https://github.com/CobanAkdemirlab/NMDescapediseasegene_paper) and the protein feature viewer at (https://github.com/CobanAkdemirlab/NMDescapediseasegene_paper/tree/main/NMDesc-protein-viewer). Scripts for the length-matched repeat, homopolymer and guanine–cytosine-content analyses (**Figure S1**), for the paralog, isoform and protein-complex comparisons (**Figure S2**), and for the gene-level and variant-level mixed models of **Figures 4–7** are included in the same repository.

## Notes

### Competing Interest Statement

The authors have declared no competing interest.

### Author Declarations

We used the human genetic variation data available from ClinVar and gnomAD control databases. They are all publicly available.

### Summary of Updates

Since the original submission, the study has been substantially revised and strengthened. The variant classification was simplified from three overlapping categories (nonsense, −1 and +1 frameshift; 148 genes) to two mechanistically interpretable classes including frameshift and stopgain (134 autosomal dominant disease genes; N = 94 and N = 64, respectively), and all gene-level comparisons were repeated against a rigorous 1:1 length-matched control set (matched on coding-sequence length and NMD-escape-region fraction). Under this more conservative design, several gene-level differences reported previously did not persist: NMD-escape disease genes are indistinguishable from their matched controls in interaction-network centrality, dosage sensitivity, tissue specificity, gene redundancy and protein-complex membership. The pathogenic signal instead proved to be local and region-specific: the NMD-escape regions of stopgain disease genes are selectively enriched for overlap with protein-protein interaction interfaces (log-odds ratio +1.22). We added gene-adjusted variant-level mixed models with explicit adjustment for truncation position, showing that pathogenic stopgain variants disrupt interaction interfaces and Pfam domains beyond what truncation position alone predicts, and we resolved the distinct modes of C-terminal remodelling of the two classes (frameshift alleles generate novel, phase-separation-competent tails, q = 0.028; stopgain alleles excise interaction-prone, disordered C-terminal sequence). The neurodevelopmental-disorder enrichment is now quantified against a matched expectation (35 observed versus 25 expected; odds ratio 1.6; P = 0.013). Most importantly, we moved beyond computational prediction by adding experimental validation: an NMD-escape truncating variant in RARB forms aberrant condensates in cells, providing direct support for the phase-separation mechanism that previously rested on structural and biophysical modelling alone (e.g., KAT6B).

