## Supplemental Figures for "Phase Separation Contributes to Pathogenicity for Nonsense Mediated Decay-Escaping Variant Alleles"

*: Co-first authors

### SUPPLEMENTARY FIGURES

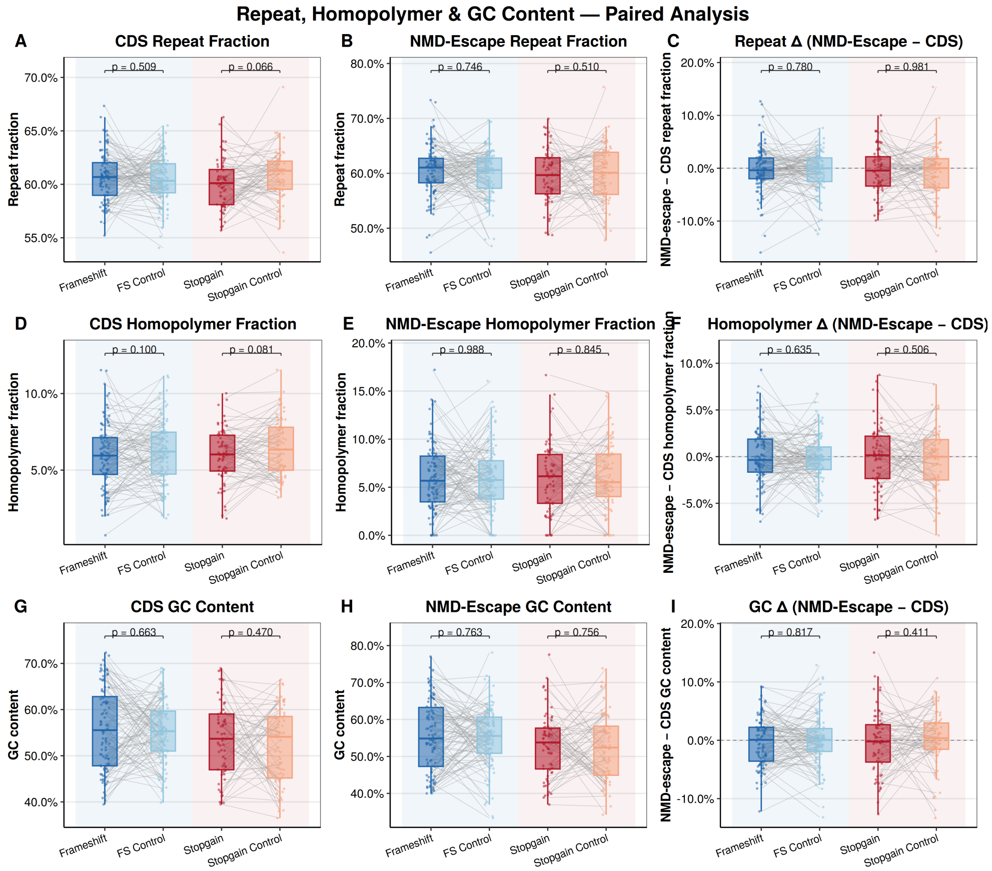


**Figure S1**. **Repeat, homopolymer and GCcontent of NMD-escape disease genes and their length-matched controls**. Paired comparisons for frameshift (versus FS control; n = 94 pairs) and stopgain (versus stopgain control; n = 64 pairs) disease genes. **(A–C)** Repeat-element fraction of the full coding sequence **(A)** and of the NMD-escape region **(B)**, and the within-transcript difference (NMD-escape − coding sequence; C). **(D–F)** The same measures for homopolymer runs. **(G–I)** GCcontent of the full coding sequence **(G)** and of the NMD-escape region **(H)**, and the within-transcript difference **(I)**. No measure differed between disease genes and their controls in either class (all P > 0.05). Boxplots show median and IQR; grey lines connect matched pairs. P values are from paired Wilcoxon signed-rank tests, Benjamini–Hochberg-adjusted within metric. NMD, nonsense-mediated decay; FS, frameshift; IQR, interquartile range.

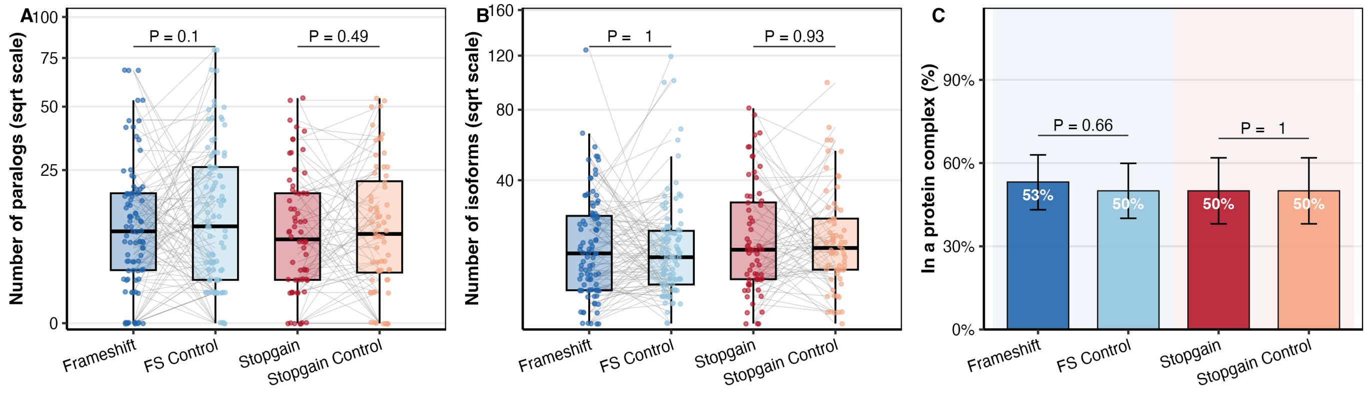


**Figure S2**. **Paralog, isoform and protein-complex counts of NMD-escape disease genes and their length-matched controls**. **(A)** Paralogs per gene (Ensembl); frameshift P = 0.1, stopgain P = 0.49. **(B)** Protein-coding transcript isoforms per gene (Ensembl); frameshift P = 1, stopgain P = 0.93. **(C)** Fraction of proteins that are a subunit of at least one human CORUM complex (bars, Wilson 95% confidence intervals), compared between disease genes and their matched controls; frameshift P = 0.66, stopgain P = 1. In A and B, counts are shown on a square-root scale, boxplots show median and IQR, and grey lines connect matched pairs. Paired Wilcoxon signed-rank test **(A, B)** or Mann–Whitney U test **(C)**. NMD, nonsense-mediated decay; IQR, interquartile range.

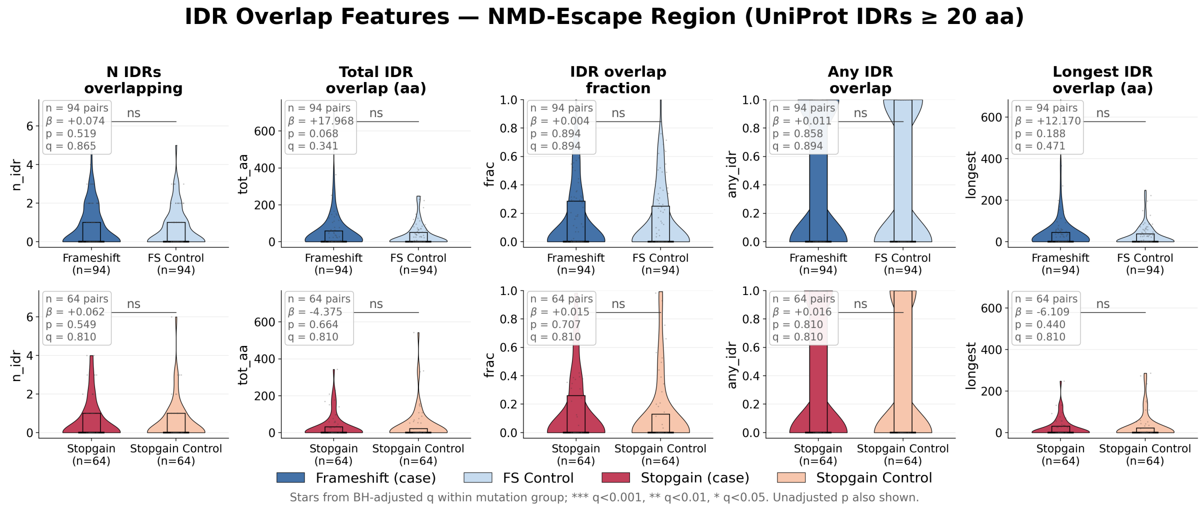


**Figure S3**. **Overlap of NMD-escape regions with intrinsically disordered regions in disease genes and their length-matched controls**. Five intrinsically disordered region-overlap metrics

(UniProt intrinsically disordered regions of at least 20 amino acids; genes absent from the intrinsically disordered region database set to zero) were computed for the NMD-escape region of each disease gene and its 1:1 length-matched control. The four count and extent metrics were tested with a linear mixed model (outcome ~ is_case + (1 | pair_id)), and the binary any-overlap metric with conditional logistic regression (is_disease ~ flag + strata(pair_id), equivalent to McNemar’s test); P values were Benjamini–Hochberg-corrected across the five metrics within each class. Top row, frameshift pairs (n = 94); bottom row, stopgain pairs (n = 64). Columns, left to right: number of overlapping intrinsically disordered regions, total overlap (amino acids), overlap fraction, any overlap, and longest single overlap (amino acids); boxes show median and IQR within each violin. None of the five metrics differed between disease genes and their controls after correction in either class (any-overlap log-odds ratio +0.065 in the frameshift class and +0.118 in the stopgain class, both q > 0.8; the count and extent metrics all non-significant). The any-overlap metric corresponds to the intrinsically disordered region flag in **Figure 4**. Abbreviations: NMD, nonsense-mediated decay; IQR, interquartile range.

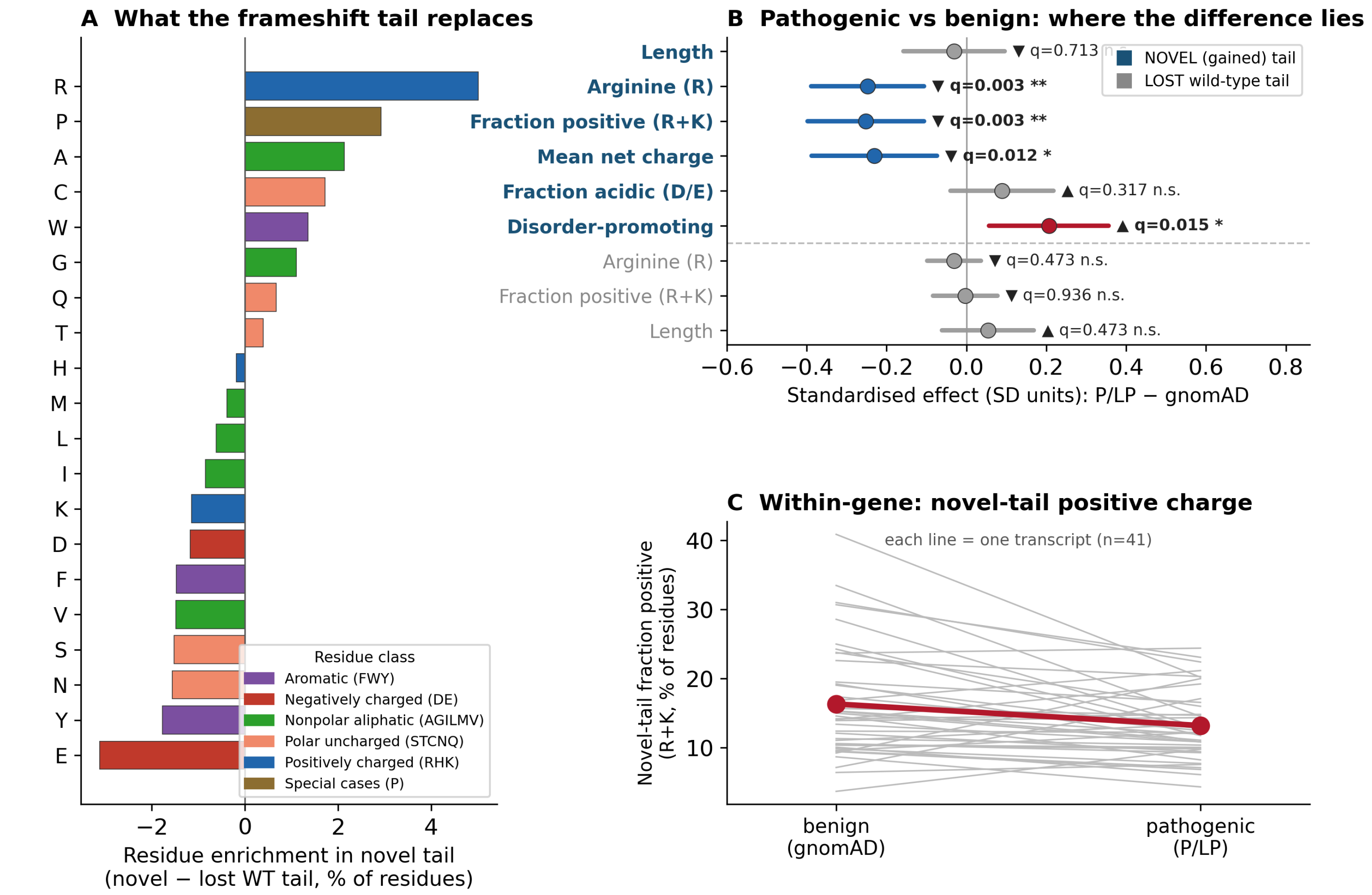


**Figure S4**. **Sequence lost and gained in frameshift NMD-escape tails**. A frameshift replaces the wild-type C-terminal sequence downstream of the variant (the 'lost' tail) with a novel out-of-frame tail translated up to the next premature stop (the 'gained' tail). For each frameshift variant the two segments were characterised and compared between pathogenic (ClinVar pathogenic or likely pathogenic) and benign (gnomAD) variants within transcripts carrying both, restricting to segments of at least 20 amino acids (41 shared transcripts; 418 pathogenic or likely pathogenic and 276 gnomAD variants). **(A)** Mean amino-acid composition of the novel (gained) tail relative to the wild-type tail it replaces (novel − lost, in percentage points, pooled across cohorts), coloured by physicochemical class. Novel out-of-frame tails are strongly enriched in arginine and proline and depleted in glutamate, tyrosine, asparagine and serine, reflecting the codon composition of the shifted reading frame. **(B)** Standardised effect (standard-deviation units) of pathogenicity (pathogenic or likely pathogenic − gnomAD) on properties of the novel (gained) tail and of the lost wild-type tail, from a linear mixed model (value ~ cohort + (1 | transcript)) with Benjamini–Hochberg correction. Within the same transcript, pathogenic variants produced novel tails with less arginine ($q = 0.003$), a lower fraction of positively charged residues (R+K; $q = 0.003$) and a lower mean net charge ($q = 0.012$) and a higher fraction of disorder-promoting residues ($q = 0.015$), whereas novel-tail length and every property of the replaced wild-type tail did not differ significantly. **(C)** Within-transcript novel-tail fraction of positively charged residues (R+K) for benign versus pathogenic variants; each grey line is one transcript and the red line the mean, showing the within-gene reduction. The reduced positive charge of pathogenic frameshift tails (**Figure 6**) therefore originates in the character of the novel out-of-frame sequence, less arginine-rich and more disorder-promoting, rather than in which wild-type sequence is lost. NMD, nonsense-mediated decay; R+K, arginine plus lysine.

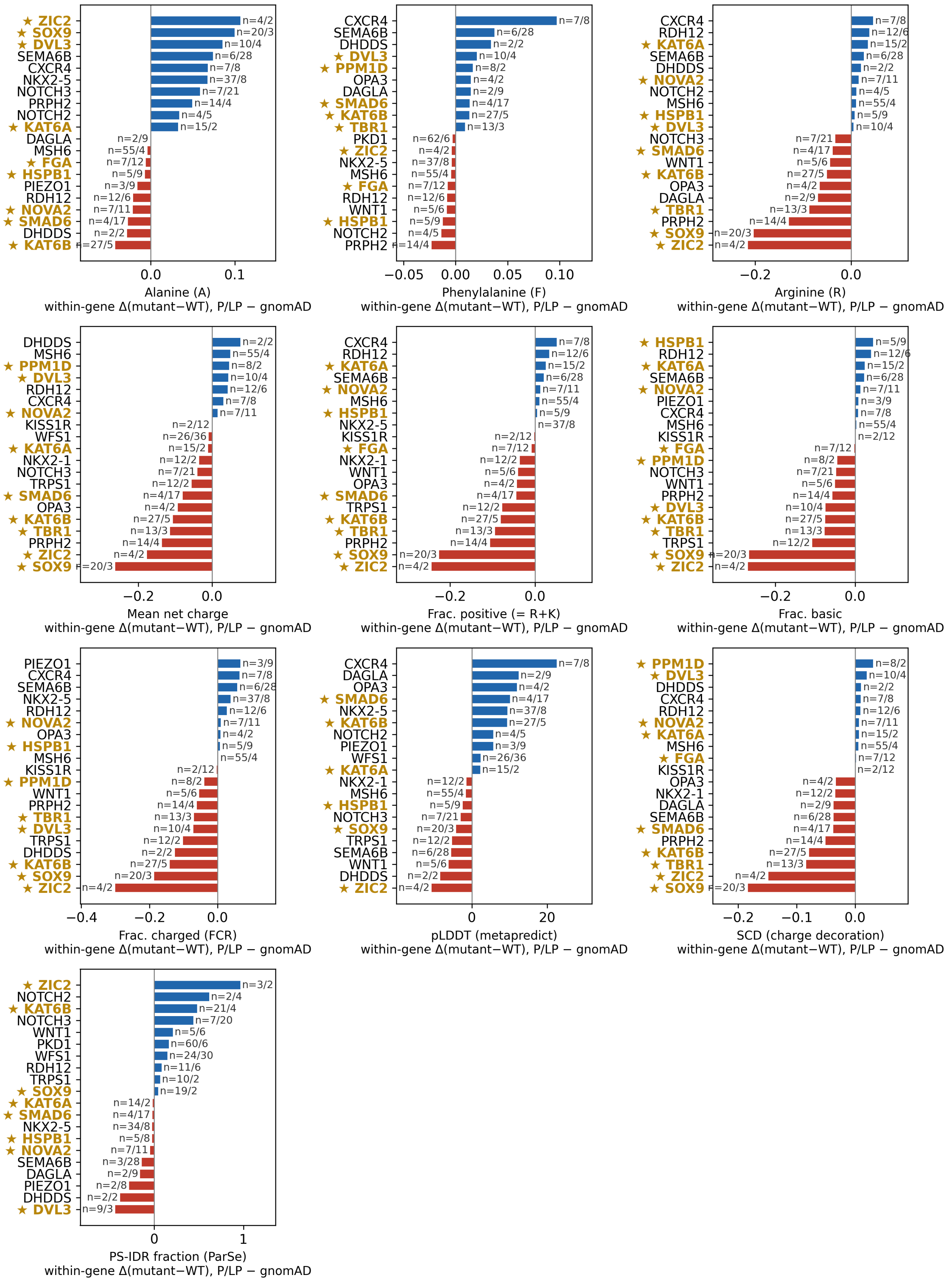
★ ZIC2
★ SOX9
★ DVL3
SEMA6B
CXCR4
NKX2-5
NOTCH3
PRPH2
NOTCH2
★ KAT6A
DAGLA
MSH6
★ FGA
★ HSPB1
PIEZO1
RDH12
★ NOVA2
★ SMAD6
DHDDS
★ KAT6B
n=4/2
n=20/3
n=10/4
n=6/28
n=7/8
n=37/8
n=7/21
n=14/4
n=4/5
n=15/2
n=2/9
n=55/4
n=7/12
n=5/9
n=3/9
n=12/6
n=7/11
n=4/17
n=2/2
n=27/5
0.0
0.1
Alanine (A)
within-gene Δ(mutant−WT), P/LP − gnomAD
CXCR4
SEMA6B
DHDDS
★ DVL3
★ PPM1D
OPA3
DAGLA
★ SMAD6
★ KAT6B
★ TBR1
PKD1
★ ZIC2
NKX2-5
MSH6
★ FGA
RDH12
WNT1
★ HSPB1
NOTCH2
PRPH2
n=7/8
n=6/28
n=2/2
n=10/4
n=8/2
n=4/2
n=2/9
n=4/17
n=27/5
n=13/3
n=62/6
n=4/2
n=37/8
n=55/4
n=7/12
n=12/6
n=5/6
n=5/9
n=4/5
n=14/4
−0.05
0.00
0.05
0.10
Phenylalanine (F)
within-gene Δ(mutant−WT), P/LP − gnomAD
CXCR4
RDH12
★ KAT6A
SEMA6B
DHDDS
★ NOVA2
NOTCH2
MSH6
★ HSPB1
★ DVL3
NOTCH3
★ SMAD6
WNT1
★ KAT6B
OPA3
DAGLA
★ TBR1
PRPH2
★ SOX9
★ ZIC2
n=7/8
n=12/6
n=15/2
n=6/28
n=2/2
n=7/11
n=4/5
n=55/4
n=5/9
n=10/4
n=7/21
n=4/17
n=5/6
n=27/5
n=4/2
n=2/9
n=13/3
n=14/4
n=20/3
n=4/2
−0.2
0.0
Arginine (R)
within-gene Δ(mutant−WT), P/LP − gnomAD
DHDDS
MSH6
★ PPM1D
★ DVL3
RDH12
CXCR4
★ NOVA2
KISS1R
WFS1
★ KAT6A
NKX2-1
NOTCH3
TRPS1
★ SMAD6
OPA3
★ KAT6B
★ TBR1
PRPH2
★ ZIC2
★ SOX9
n=2/2
n=55/4
n=8/2
n=10/4
n=12/6
n=7/8
n=7/11
n=2/12
n=26/36
n=15/2
n=12/2
n=7/21
n=12/2
n=4/17
n=4/2
n=27/5
n=13/3
n=14/4
n=4/2
n=20/3
−0.2
0.0
Mean net charge
within-gene Δ(mutant−WT), P/LP − gnomAD
CXCR4
RDH12
★ KAT6A
SEMA6B
★ NOVA2
MSH6
★ HSPB1
NKX2-5
KISS1R
★ FGA
NKX2-1
WNT1
OPA3
★ SMAD6
TRPS1
★ KAT6B
★ TBR1
PRPH2
★ SOX9
★ ZIC2
n=7/8
n=12/6
n=15/2
n=6/28
n=7/11
n=55/4
n=5/9
n=37/8
n=2/12
n=7/12
n=12/2
n=5/6
n=4/2
n=4/17
n=12/2
n=27/5
n=13/3
n=14/4
n=20/3
n=4/2
−0.2
0.0
Frac. positive (= R+K)
within-gene Δ(mutant−WT), P/LP − gnomAD
★ HSPB1
RDH12
★ KAT6A
SEMA6B
★ NOVA2
PIEZO1
CXCR4
MSH6
KISS1R
★ FGA
★ PPM1D
NOTCH3
WNT1
PRPH2
★ DVL3
★ KAT6B
★ TBR1
TRPS1
★ SOX9
★ ZIC2
n=5/9
n=12/6
n=15/2
n=6/28
n=7/11
n=3/9
n=7/8
n=55/4
n=2/12
n=7/12
n=8/2
n=7/21
n=5/6
n=14/4
n=10/4
n=27/5
n=13/3
n=12/2
n=20/3
n=4/2
−0.2
0.0
Frac. basic
within-gene Δ(mutant−WT), P/LP − gnomAD
PIEZO1
CXCR4
SEMA6B
NKX2-5
RDH12
★ NOVA2
OPA3
★ HSPB1
MSH6
KISS1R
★ PPM1D
WNT1
PRPH2
★ TBR1
★ DVL3
TRPS1
DHDDS
★ KAT6B
★ SOX9
★ ZIC2
n=3/9
n=7/8
n=6/28
n=37/8
n=12/6
n=7/11
n=4/2
n=5/9
n=55/4
n=2/12
n=8/2
n=5/6
n=14/4
n=13/3
n=10/4
n=12/2
n=2/2
n=27/5
n=20/3
n=4/2
−0.4
−0.2
0.0
Frac. charged (FCR)
within-gene Δ(mutant−WT), P/LP − gnomAD
CXCR4
DAGLA
OPA3
★ SMAD6
NKX2-5
★ KAT6B
NOTCH2
PIEZO1
WFS1
★ KAT6A
NKX2-1
MSH6
★ HSPB1
NOTCH3
★ SOX9
TRPS1
SEMA6B
WNT1
DHDDS
★ ZIC2
n=7/8
n=2/9
n=4/2
n=4/17
n=37/8
n=27/5
n=4/5
n=3/9
n=26/36
n=15/2
n=12/2
n=55/4
n=5/9
n=7/21
n=20/3
n=12/2
n=6/28
n=5/6
n=2/2
n=4/2
0
20
pLDDT (metapredict)
within-gene Δ(mutant−WT), P/LP − gnomAD
★ PPM1D
★ DVL3
DHDDS
CXCR4
RDH12
★ NOVA2
★ KAT6A
MSH6
★ FGA
KISS1R
OPA3
NKX2-1
DAGLA
SEMA6B
★ SMAD6
PRPH2
★ KAT6B
★ TBR1
★ ZIC2
★ SOX9
n=8/2
n=10/4
n=2/2
n=7/8
n=12/6
n=7/11
n=15/2
n=55/4
n=7/12
n=2/12
n=4/2
n=12/2
n=2/9
n=6/28
n=4/17
n=14/4
n=27/5
n=13/3
n=4/2
n=20/3
−0.2
−0.1
0.0
SCD (charge decoration)
within-gene Δ(mutant−WT), P/LP − gnomAD
★ ZIC2
NOTCH2
★ KAT6B
NOTCH3
WNT1
PKD1
WFS1
RDH12
TRPS1
★ SOX9
★ KAT6A
★ SMAD6
NKX2-5
★ HSPB1
★ NOVA2
SEMA6B
DAGLA
PIEZO1
DHDDS
★ DVL3
n=3/2
n=2/4
n=21/4
n=7/20
n=5/6
n=60/6
n=24/30
n=11/6
n=10/2
n=19/2
n=14/2
n=4/17
n=34/8
n=5/8
n=7/11
n=3/28
n=2/9
n=2/8
n=2/2
n=9/3
0
1
PS-IDR fraction (ParSe)
within-gene Δ(mutant−WT), P/LP − gnomAD

**Figure S5**. **Gene-level contributions to the frameshift escape-tail composition effects**. Per-gene ranking of the within-gene pathogenic−benign difference for the features that differ significantly in **Figure 6**: alanine (A), phenylalanine (F) and arginine (R) content (**Figure 6A**); mean net charge, fraction positive (= R+K), fraction basic, fraction charged (fraction of charged residues) and pLDDT (**Figure 6C**); charge decoration (sequence charge decoration, **Figure 6D**); and the ParSe phase-separating intrinsically disordered region fraction (**Figure 6E**). Bars show the within-gene difference between pathogenic and benign variants (pathogenic or likely pathogenic − gnomAD) in the change relative to wild-type on the post-frameshift tail (Δ = mutant − wild-type). For the composition and charge features, only tails of at least 20 amino acids and genes with at least two variants in each cohort are included (30 genes; 395 pathogenic or likely pathogenic and 246 gnomAD variants); the phase-separating intrinsically disordered region panel uses the ParSe escape tails of at least 25 amino acids on the same basis (28 genes; 344 pathogenic or likely pathogenic and 217 gnomAD variants). The ten genes with the most positive and the ten with the most negative effects are shown for each feature (blue, higher in pathogenic; red, lower); n = pathogenic or likely pathogenic/gnomAD variants per gene. Gold stars mark genes encoding known phase-separation proteins. The phase-separation proteins SOX9, ZIC2 and KAT6B are among the genes with the largest loss of positive and total charge in pathogenic tails, so the aggregate effects in **Figure 6** reflect many genes rather than a single outlier. pLDDT, predicted local distance difference test; R+K, arginine plus lysine.

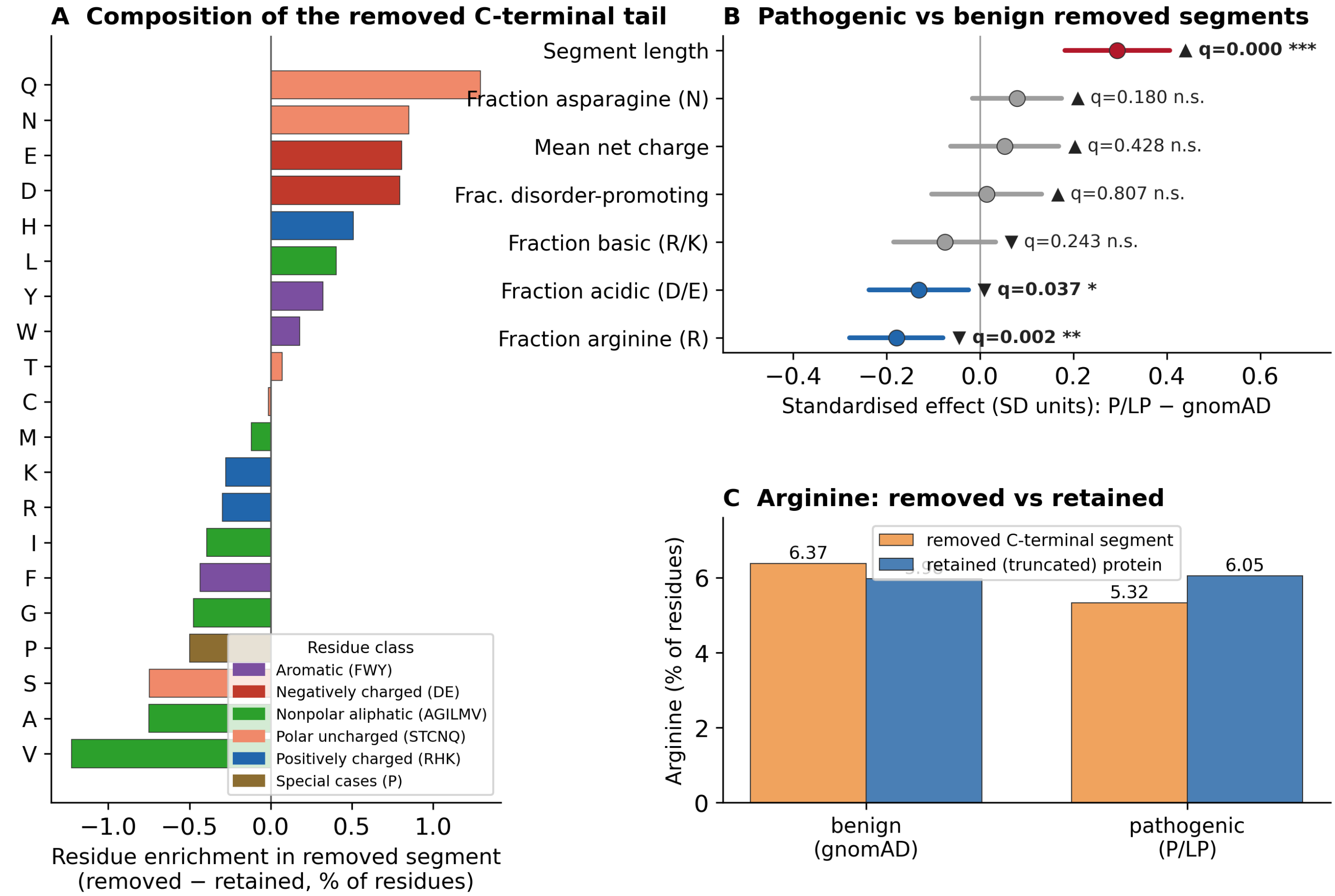


**Figure S6**. **Composition of the C-terminal segments removed by stopgain truncations**. For each stopgain variant, the C-terminal segment removed by the premature stop (the wild-type residues downstream of the truncation) was characterised and compared between pathogenic (ClinVar pathogenic or likely pathogenic) and benign (gnomAD) variants within transcripts carrying both (47 transcripts; 337 pathogenic or likely pathogenic and 210 gnomAD variants). **(A)** Amino-acid composition of the removed C-terminal tail relative to the retained (truncated) protein (removed − retained, in percentage points, pooled across cohorts), coloured by physicochemical class. The removed tails are enriched in polar and acidic residues (Q, N, E, D) and depleted in arginine and other residues that concentrate in the ordered core, so truncation raises the arginine and basic-residue fraction of the retained protein. **(B)** Standardised effect (standard-deviation units) of pathogenicity (pathogenic or likely pathogenic − gnomAD) on properties of the removed segment, from a linear mixed model (value ~ cohort + (1 | transcript)) with Benjamini–Hochberg correction. Pathogenic truncations remove much longer segments ($q < 0.001$) that are more arginine-depleted ($q = 0.002$) and modestly less acidic ($q = 0.037$); the other properties do not differ significantly. **(C)** Mean arginine content of the removed segment versus the retained protein, by cohort. Benign truncations remove slightly arginine-rich C-terminal tails (removed > retained), which lowers arginine in the product, whereas pathogenic truncations remove longer, arginine-poor segments (removed < retained), concentrating arginine in the retained protein, which is correspondingly more arginine-dense in pathogenic variants (pathogenic or likely pathogenic − gnomAD, $P = 0.0004$). The higher arginine, basic-residue and net-positive-charge content of pathogenic stopgain products (**Figure 7**) therefore reflects loss of long, arginine-poor, polar and disorder-promoting C-terminal sequence rather than retention of arginine-rich low-complexity regions. NMD, nonsense-mediated decay.

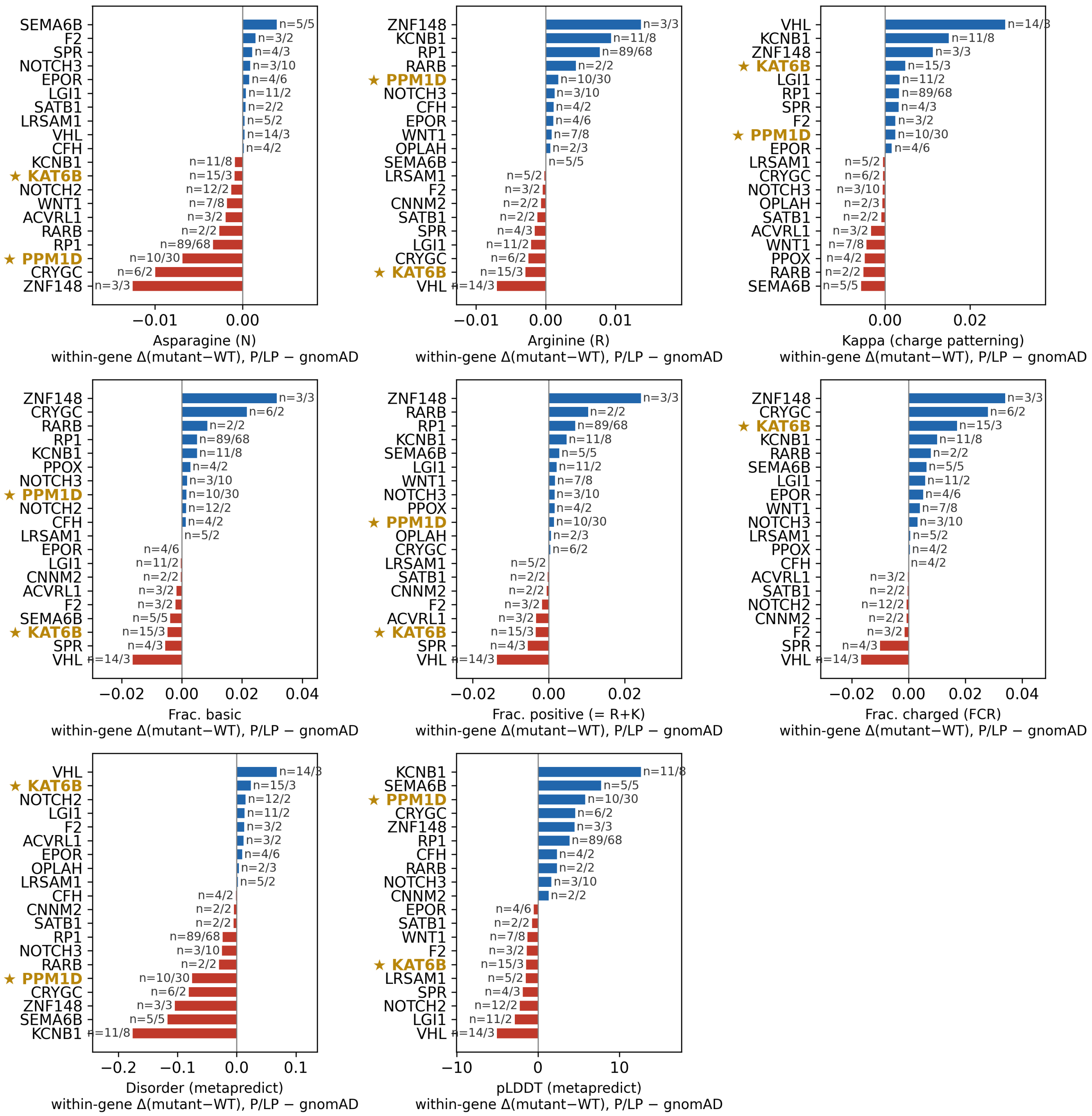


**Figure S7**. **Gene-level contributions to the stopgain whole-protein composition effects**. As in **Figure S5**, but for stopgain variants and the features that differ significantly in **Figure 7**: asparagine and arginine content (**Figure 7A**), charge patterning (κ), fraction basic, fraction positive (R+K), fraction charged (fraction of charged residues), predicted disorder and pLDDT (**Figure 7C**). Bars show the within-gene difference between pathogenic and benign variants (pathogenic or likely pathogenic − gnomAD) in the change between the truncated mutant and the full-length wild-type protein (ΔFull = mutant − wild-type). Genes with at least two variants in each cohort are included (23 genes; 221 pathogenic or likely pathogenic and 172 gnomAD variants), and the ten genes with the most positive and the ten with the most negative effects are shown for each feature (blue, higher in pathogenic; red, lower); n = pathogenic or likely pathogenic/gnomAD variants per gene. Gold stars mark genes encoding known phase-separation proteins. Pathogenic truncations in KCNB1, SEMA6B, ZNF148 and CRYGC remove the most disordered sequence, whereas VHL and KAT6B shift in the opposite direction, so the whole-protein effects in **Figure 7** are spread across many genes rather than driven by one. κ, the Das–Pappu charge-patterning parameter; R+K, arginine plus lysine; pLDDT, predicted local distance difference test.

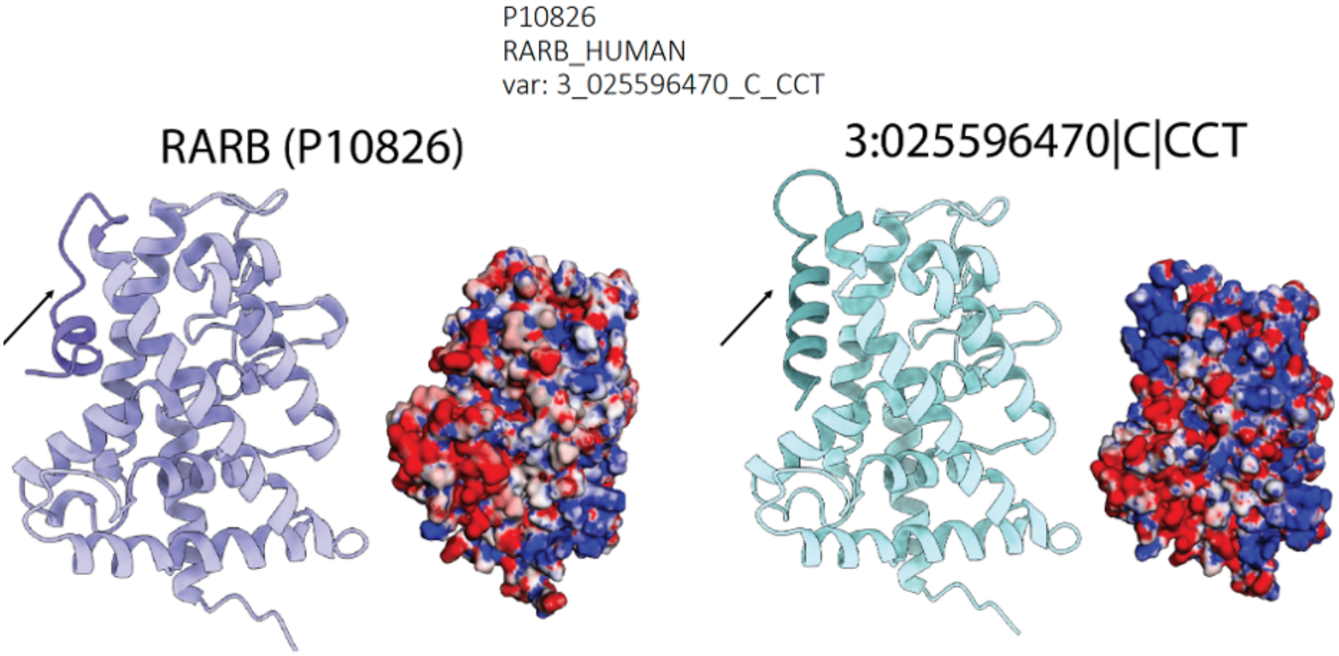


**Figure S8**. **Ribbon and surface electrostatic representations illustrate the predicted structural consequences of a pathogenic NMD-escape frameshift variant in *RARB* (P10826)**. Left: Wild-type RARB structure shown as a ribbon diagram and corresponding electrostatic surface map, highlighting the canonical C-terminal ligand-binding domain. Right: Predicted structure for the frameshift variant 3:025596470|C|CCT, which introduces an alternative

C-terminal truncation and/or alteration. Ribbon and electrostatic surface views reveal substantial alterations in C-terminal sequence and charge distribution, including loss of a disordered, highly charged tail. These structural disruptions may perturb RARB protein stability, protein–protein interactions, or nuclear receptor regulatory function, providing a potential mechanism linking NMD-escape to disease. NMD, nonsense-mediated decay.
